# Co-development of anxiety and depression in UK and Brazil youth; a cross-country comparison

**DOI:** 10.64898/2026.06.22.26356231

**Authors:** Amy Shakeshaft, Lucy Barrass, Bushra Farooq, Lucy Riglin, Ana Goncalves Soares, Hannah J. Jones, Nicky Lidbetter, Duleeka J. Knipe, Ian Penton-Voak, Marina X. Carpena, Iná S. Santos, Luciana Tovo-Rodrigues, Jon Heron, Frances Rice, Alicia Matijasevic, Laura D. Howe

**Affiliations:** Population Health Sciences, Bristol Medical School, University of Bristol, Bristol, UK; Wolfson Centre for Young People’s Mental Health, Division of Psychological Medicine and Clinical Neuroscience, School of Medicine, Cardiff University, Cardiff, UK; Anxiety UK, Manchester, UK; School of Psychological Science, University of Bristol, Bristol, UK; NIHR Biomedical Research Centre at the University Hospitals Bristol NHS Foundation Trust, Bristol, UK; Postgraduate Program in Epidemiology, Federal University of Pelotas, Pelotas, Rio Grande do Sul, Brazil; Departamento de Medicina Preventiva, Faculdade de Medicina FMUSP, Universidade de São Paulo, São Paulo, Brazil

**Keywords:** ALSPAC, Pelotas birth cohort, Co-development, Cross-context, Developmental trajectories, Anxiety, Depression, Emotional problems, Youth mental health, Global mental health

## Abstract

**Importance:** Anxiety and depression frequently co-occur and show developmentally patterned co-development from childhood to adolescence. Adult psychiatric outcomes vary according to the timing, sequencing, and persistence of early symptoms, yet it remains unclear whether patterns of co-development are comparable across high-income and low– and middle-income country contexts.

**Objective:** Examine joint developmental trajectories of anxiety and depression from childhood to adolescence and their associations with anxiety and depression diagnoses in young adulthood.

**Design, Setting and Participants:** Population-based prospective cohort studies in the UK (Avon Longitudinal Study of Parents and Children [ALSPAC], N=9,586) and Brazil (Pelotas 2004 Birth Cohort, N=3,815).

**Main Outcomes and Measures:** Trajectories were derived using parallel-process latent growth models and latent class growth analyses of anxiety and depression using the Development and Well-Being Assessment at early childhood (6–7 years), middle childhood (10–11 years), and adolescence (13–15 years). Diagnoses of anxiety and depression at 18 years were assessed via the Clinical Interview Schedule (ALSPAC) and the Mini International Neuropsychiatric Interview (Pelotas).

**Results:** Prevalence of anxiety and depression from early childhood to adolescence was similar across cohorts. Co-development was stronger in ALSPAC, with modest increases in both conditions, whereas in Pelotas, anxiety increased rapidly while depression showed little average change. In both cohorts, four trajectory classes were identified: stable-low (ALSPAC, 41%; Pelotas, 54%), increasing (31%; 28%), decreasing (23%; 15%), and persistent-high anxiety/increasing depression (5%; 3%). Compared with the stable-low class, youth in the increasing and persistent-high classes had elevated odds of depression (ALSPAC: OR=2.0 [95% CI, 1.4–2.8] and 4.2 [2.6–6.7]; Pelotas: 2.2 [1.5–3.3] and 2.9 [1.4–6.0]) and anxiety in young adulthood (ALSPAC: 1.6 [1.2–2.2] and 4.8 [3.2–7.0]; Pelotas: 1.7 [1.2–2.6] and 2.9 [1.5–5.8]). No increased risk was observed in the decreasing class.

**Conclusions and Relevance:** Patterns of anxiety and depression co-development were comparable across the UK and Brazil, suggesting shared developmental pathways. However, more rapid increases in anxiety among Brazilian youth may reflect context-specific risk factors. Persistence or emergence beyond early childhood was critical for identifying later diagnostic risk in both settings, highlighting the importance of early monitoring and intervention.

**Key points:** **Question:** How does the co-development of anxiety and depression from early childhood to adolescence differ between high-income (HICs) and low– and middle-income countries (LMICs)?

**Findings:** In this comparative longitudinal study of two population-based birth cohorts from the UK (N = 9,586) and Brazil (N = 3,815), trajectories of anxiety and depression co-development from childhood through adolescence were broadly similar across settings and were predictive of anxiety and depression diagnoses in young adulthood.

**Meaning:** Despite substantial socioeconomic and contextual differences between the UK and Brazil, developmental pathways of anxiety and depression co-development appear largely comparable, suggesting shared underlying developmental processes across these contexts.

## Introduction

Mental health problems are the leading cause of global disease burden^1^ and affect around 11% of young people between 5-24 years of age^2^, with rates increasing. Anxiety conditions, including generalized anxiety disorder (GAD), specific phobias, social anxiety and separation anxiety, and depression are common in young people and frequently co-occur^3^. They share much in common, including a female preponderance, environmental risk factors^4^, and genetic liability^5^. Developmentally, anxiety conditions generally emerge earlier than depression; appearing in childhood (e.g. separation anxiety, phobias) or adolescence (e.g. social anxiety, GAD), compared with major depression in young adulthood^6^. Anxiety and depression have short and long-term consequences for young people^7,8^. The persistence of symptoms into adulthood is common, particularly in females^9^, as is transitioning from one condition to another^10–13^, and the risk of poor outcomes is elevated in those with co-occurring depression and anxiety^4^.

However, a crucial knowledge gap remains; whether patterns of anxiety and depression co-development, that is how they develop in relation to one another over time, are consistent between high-income countries (HICs) and low– and middle-income countries (LMICs). Most studies have been carried out in HIC populations so evidence from LMICs is lacking. In LMICs, young people are exposed to greater levels of environmental determinants of poor mental health, including childhood adversity, and housing and food insecurity^14–17^ and therefore global studies are essential to ensure interventions are appropriate and effective across contexts.

Therefore, we aim to examine and compare the development of anxiety and depression in two longitudinal birth cohorts: i) the Avon Longitudinal Study of Parents and Children (UK; HIC), and ii) the Pelotas 2004 Cohort (Brazil; an upper-middle-income country). We analyse joint developmental trajectories of anxiety and depression from early childhood to adolescence to determine whether co-development is similar across these settings, and assess how trajectories are associated with anxiety and depression diagnoses in young adulthood.

Prior cross-sectional analyses in UK and Brazilian birth cohorts from 1990s-2000s suggest that, on average, youth in Brazil experience higher levels of emotional problems than their UK counterparts^18^, and that there are considerable social differences, including elevated rates of childhood adversity and parental mental health problems in Brazil^19^. Therefore, we hypothesize that patterns of anxiety and depression co-development may differ across contexts, with young people in Brazil being less likely to exhibit a consistently low anxiety/depression profile. Further, we hypothesize that, in both cohorts, those with persistent anxiety/depression profile between early childhood and adolescence will show the highest odds of anxiety and depression diagnoses in young adulthood.

## Methods

This study has been preregistered at https://osf.io/x9epn/. For deviations see the **Supplement.**

### Samples

The Avon Longitudinal Study of Parents and Children (ALSPAC) is an ongoing longitudinal study which recruited pregnant women in the Avon region of South-West England, due to give birth between 1^st^ April 1991 and 31^st^ December 1992^20–22^. The core sample consisted of 14,541 mothers and of these pregnancies, 13,988 children were alive at 1 year. Following initial recruitment, an additional 913 children were recruited in three phases. Data were collected from families repeatedly and managed using REDCap^23^. The study website contains details of available data through a searchable data dictionary and variable search tool (http://www.bristol.ac.uk/alspac/researchers/our-data/). For further details, see the cohort profile^21^.

The Pelotas 2004 Birth Cohort is a population-based study that recruited babies born between 1^st^ January and 31^st^ December 2004 in the city of Pelotas, Southern Brazil. Hospitals with maternity wards were visited daily, and all live births were eligible for enrolment in the study^24,25^. 4,231 newborns were included in the cohort, representing 99.2% of all births in the city that year. All participants were assessed at birth and followed up until young adulthood. For further details, see the cohort profiles^24,25^.

For both cohorts, the study sample consisted of participants with at least one complete measure of anxiety or depression from at least one timepoint (early childhood, middle childhood or adolescence).

See the **Supplement** for details of ethical approvals.

### Measures

#### Anxiety and depression

We used the Development and Wellbeing Assessment^26^ (DAWBA), a structured research diagnostic interview, consistent with DSM (Diagnostic and Statistical Manual of Mental Disorders) and ICD (International Classification of Disease), to capture major depressive disorder (MDD) and anxiety conditions. The DAWBA has been translated and validated for use in Brazilian populations^27^. In both cohorts, the DAWBA was parent-reported in early childhood, middle childhood, and adolescence (ALSPAC ages 7, 10 and 13 years; Pelotas ages 6, 11 and 15 years). We used DAWBA band variables for any anxiety diagnosis (including GAD, separation anxiety, specific phobias, social anxiety) and depression diagnosis. These bands use symptoms and impact to generate ordered-categorical measures indicating the probability that an individual has the condition (<0.1%, 0.5%, 3%, 15%, 50%, >70%). The DAWBA bands are validated in British and Norwegian samples^28^. We also use DAWBA-derived diagnoses based on those >50% likelihood of diagnosis^26^.

#### Anxiety and depression in young adulthood

In ALSPAC, depression and anxiety were assessed at age 18 using a self-administered revised Clinical Interview Schedule (CIS-R) questionnaire^29^. The CIS-R assigns ICD-10 diagnoses of depression and the presence of symptoms of anxiety conditions^30^. In Pelotas, the Mini International Neuropsychiatric Interview (MINI) was used to assess depression and any anxiety condition at age 18. The MINI is a brief, structured diagnostic interview assessing psychiatric conditions, validated for use in Brazil^31^.

### Statistical analysis

Results are presented in line with the Guidelines for Reporting on Latent Trajectory Studies (GRoLTS) checklist^32^. Analysis scripts are available in the **Supplement.**

Descriptive statistics of the prevalence of depression and anxiety conditions at each developmental stage are presented in both cohorts and compared using chi-squared tests. Cross-sectional correlations between DAWBA bands for all anxiety conditions and depression were tested using Spearman’s rank.

#### Growth curve and mixture modelling

Parallel-process latent growth models (LGMs) were fitted in each cohort to examine the co-development of anxiety and depression, using growth curve modelling for ordered-categorical data in MPlus^33^, using Weighted Least Squares Mean and Variance adjusted (WLSMV) estimator. Anxiety and depression intercepts were specified at the youngest age (ALSPAC age 7, Pelotas age 6) and slopes capture linear change across the study period. As parent-reported data were only available at three timepoints, we were unable to model non-linear change. For details of model specification see the **Supplement.**

We examined heterogeneity in the co-development of anxiety and depression using latent class growth analysis (LCGA), using maximum likelihood (ML) estimator as mixture modelling is not compatible with WLSMV estimator. Whereas LGM summarize average developmental change, LCGA identifies subgroups within the population that follow different developmental trajectories. Starting with a single k-class model, k+1 solutions were fitted until the optimal model was reached, as determined using Lo-Mendell-Rubin (LMR) tests, Akaike information criterion (AIC), Bayesian information criterion (BIC), smallest class size information, and the plausibility and interpretability of classes. Other model fit measures, including log-likelihood, entropy and Bootstrap Likelihood Ratio Test (BLRT), are reported in the **Supplement.**

We then used the manual bias-adjusted three-step approach^34–36^ to examine associations between trajectory classes and anxiety and depression diagnoses in young adulthood, adjusting for uncertainty in class assignment.

#### Missing data

Missing data for each analysis variable are presented in **eTables 1–2**. LCGA models were estimated using full information maximum likelihood (FIML) to handle missing data in DAWBA assessments, as supported in MPlus. For LGM, which used WLSMV estimator and so not compatible with FIML, multiple imputation by chained equations (MICE, 200 imputations; 25 iterations) was used to impute missing anxiety and depression data. Similarly, MICE was also used to impute missingness in anxiety and depression diagnoses when testing associations between trajectory classes and young adulthood diagnoses. For these analyses, estimates pooled across imputed datasets. For further details, see the **Supplement.**

### Secondary and sensitivity analyses

These analyses are presented in the **Supplement**. We examined the prevalence of each anxiety condition in each trajectory class in both cohorts. Since persistence of depression/anxiety is more commonly observed in females, we investigated whether patterns of anxiety and depression development differ by sex in both cohorts by performing sex-stratified parallel-process LCGA. Finally, we present descriptive statistics from the unimputed samples.

## Results

The final study sample comprised N=9,586 in ALSPAC (49% female) and N=3,815 in Pelotas (48% female). The prevalence of any anxiety condition and depression were similar in both cohorts during early childhood and adolescence, but during middle childhood there were higher rates of depression in ALSPAC (1.1% vs 0.4%) and of anxiety in Pelotas (3.6% vs. 2.8%, **Table 1**). Developmental patterns of anxiety and depression prevalence were as expected and similar in both cohorts. Depression prevalence was highest in adolescence, as was social anxiety and GAD (although rates of GAD were very low in Pelotas). The prevalence of any anxiety condition, separation anxiety and specific phobias peaked in middle childhood in both cohorts. For prevalence rates in young adulthood, see **eTable 3**.

**Table 1.** – Anxiety and depression diagnoses across early childhood, middle childhood and adolescence in ALSPAC and Pelotas 2004 birth cohorts. Diagnoses derived from the DAWBA.

| Diagnosis | Developmental stage* | ALSPAC<br>(total N = 9,586) |  | Pelotas<br>(total N = 3,815) |  | Chi-sq<br>p-value |
| --- | --- | --- | --- | --- | --- | --- |
|  |  | N | % | N | % |  |
| Depression | Early childhood | 62 | 0.7 | 18 | 0.5 | 0.29 |
|  | Middle childhood | 108 | 1.1 | 15 | 0.4 | 8.9x10<sup>-5</sup> |
|  | Adolescence | 119 | 1.2 | 57 | 1.5 | 0.28 |
| Any anxiety condition | Early childhood | 190 | 2.0 | 84 | 2.2 | 0.46 |
|  | Middle childhood | 273 | 2.8 | 137 | 3.6 | 0.03 |
|  | Adolescence | 229 | 2.4 | 75 | 2.0 | 0.16 |
| Specific phobia | Early childhood | 108 | 1.1 | 52 | 1.4 | 0.29 |
|  | Middle childhood | 118 | 1.2 | 95 | 2.5 | 2.2x10<sup>-7</sup> |
|  | Adolescence | 58 | 0.6 | 58 | 1.5 | 4.2x10<sup>-7</sup> |
| Social anxiety | Early childhood | 11 | 0.1 | <5<sup>^</sup> | <0.1 | 0.46 |
|  | Middle childhood | 39 | 0.4 | <5<sup>^</sup> | <0.1 | 0.004 |
|  | Adolescence | 56 | 0.6 | 13 | 0.3 | 0.10 |
| GAD | Early childhood | 20 | 0.2 | 0 | 0 | 0.01 |
|  | Middle childhood | 58 | 0.6 | 5 | 0.1 | 0.0005 |
|  | Adolescence | 69 | 0.7 | 5 | 0.1 | 5.8x10<sup>-5</sup> |
| Separation anxiety | Early childhood | 85 | 0.9 | 35 | 0.9 | 0.94 |
|  | Middle childhood | 131 | 1.4 | 42 | 1.1 | 0.25 |
|  | Adolescence | 115 | 1.2 | 8 | 0.2 | 1.0x10<sup>-7</sup> |
| Anxiety depression comorbidity | Early childhood | 22 | 0.2 | 10 | 0.3 | 0.88 |
|  | Middle childhood | 43 | 0.4 | 7 | 0.2 | 0.03 |
|  | Adolescence | 52 | 0.5 | 8 | 0.2 | 0.01 |

\*ALSPAC early childhood: 7 years, middle childhood: 10 years, adolescence: 13 years. Pelotas 2004 early childhood: 6 years, middle childhood: 11 years, adolescence: 15 years.
<sup>^</sup>Due to disclosure protocols, cell sizes of <5 are not given as exact values.

**ETable 4** shows distributions of DAWBA bands at each assessment age. Correlations between any anxiety condition and depression DAWBA bands were moderate (r=0.3) and similar in both cohorts, and at each developmental stage (see **eFigure 1** for all correlations).

**Figure 1.**
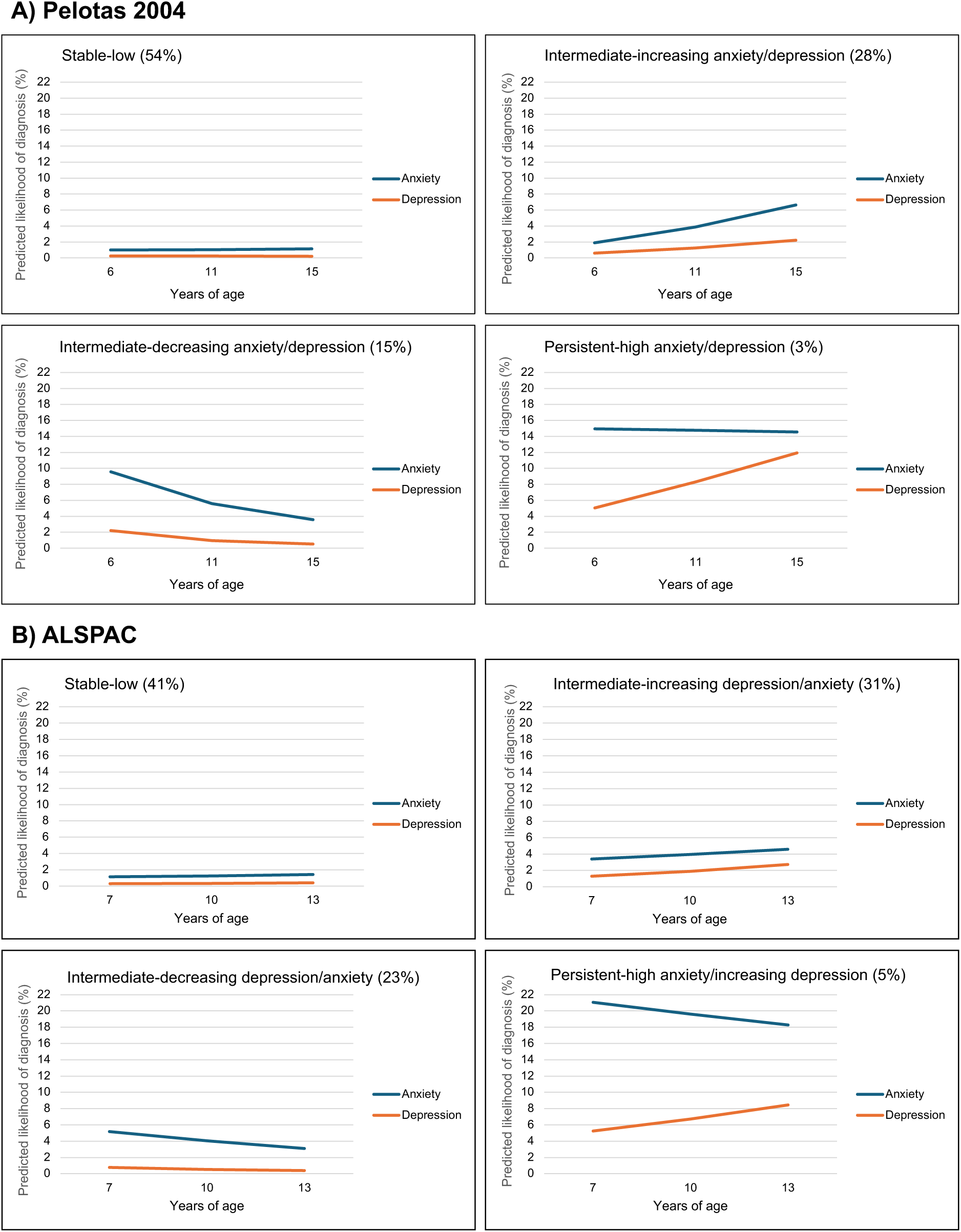
– 4-Class LCGA solution for anxiety and depression in A) Pelotas 2004 and B) ALSPAC.

### Trajectories of anxiety and depression in ALSPAC and Pelotas 2004

**ETable 5** shows parameter estimates from parallel-process LGM in both cohorts. In ALSPAC, on average, the likelihood of depression diagnosis (slope=0.016, p<0.001) and any anxiety condition diagnosis increased (slope=0.005, p=0.048) between ages 7-13 years. Baseline anxiety and depression were strongly correlated (intercept correlation=0.70, p<0.001), as were slopes (slope correlation=0.72, p<0.001), demonstrating coupled developmental change: i.e., on average, individuals with steeper increases in anxiety showed correspondingly steeper increases in depression.

In Pelotas, on average, the likelihood of anxiety diagnosis increased between ages 6-15 years (slope=0.042, p<0.001), but an increase was not as evident for depression (slope=0.009, p=0.13). Like ALSPAC, baseline anxiety and depression were correlated (intercept correlation=0.52, p<0.001). However, there was less evidence of correlation among depression and anxiety slopes in Pelotas (slope correlation=0.88, p=0.31).

Results from anxiety-only and depression-only LCGA are presented in the **Supplement (eTables 6-9).**

Model fit statistics for parallel-process anxiety and depression LCGA are presented in **eTables 10-11**. In Pelotas, the 4-class solution showed optimal fit (**Figure 1A**); with a *stable-low* anxiety and depression class (54%), an *intermediate-increasing* anxiety and depression class (28%), an *intermediate-decreasing* anxiety and depression class (15%) and a *persistent-high* anxiety and depression class (3%, although depression appears to increase in this class, there was not strong evidence for this increase according to model estimates [slope estimate=0.129, p=0.24]; **eTable 12**).

In ALSPAC, the optimal solution was less clear, as, for 2-6 class models, the LMR p-value was ≤0.001 and smallest class size >1%. Visualization of 5– and 6-class solutions showed limited differentiation between several trajectory groups, all showing stable-low depression and anxiety. Visualization of the 4-class solution (**Figure 1B**) showed more distinct classes, like Pelotas; i) *stable-low* anxiety and depression (41%), ii) *intermediate-increasing* anxiety and depression (31%), iii) *intermediate-decreasing* anxiety and depression (23%), and iv) *persistent-high anxiety/increasing depression* (5%). For the *intermediate-decreasing* class, while both depression and anxiety decrease over time (anxiety = –0.17, p<0.001; depression = –0.19, p<0.001; **eTable 12**), depression is at a similar level to the *stable-low* class throughout.

### Associations between trajectory classes and young adult anxiety and depression diagnoses

In ALSPAC, the *intermediate-increasing* class (anxiety OR=1.63 [1.20-2.20]; depression OR=2.02 [1.44-2.82]) and *high* class (anxiety OR=4.77 [3.24-7.03]; depression OR=4.19 [2.61-6.71]) had higher odds of anxiety and depression diagnoses in young adulthood than the *stable-low* class (**Figure 2; eTable 13**). This pattern of results was repeated in Pelotas, whereby both the *intermediate-increasing* class (anxiety OR=1.72 [1.16-2.56]; depression OR=2.21 [1.46-3.33]) and *high* class (anxiety OR=2.94[1.48-5.83]; depression OR=2.87 [1.38-5.97]) had elevated odds. The *intermediate-decreasing* class did not differ from the *stable-low* class in ALSPAC (anxiety OR=1.21 [0.77-1,90]; depression OR=0.84 [0.41-1.72]) or Pelotas (anxiety OR=0.92 [0.51-1.63]; depression OR=1.50 [0.91-2.47]).

**Figure 2.**
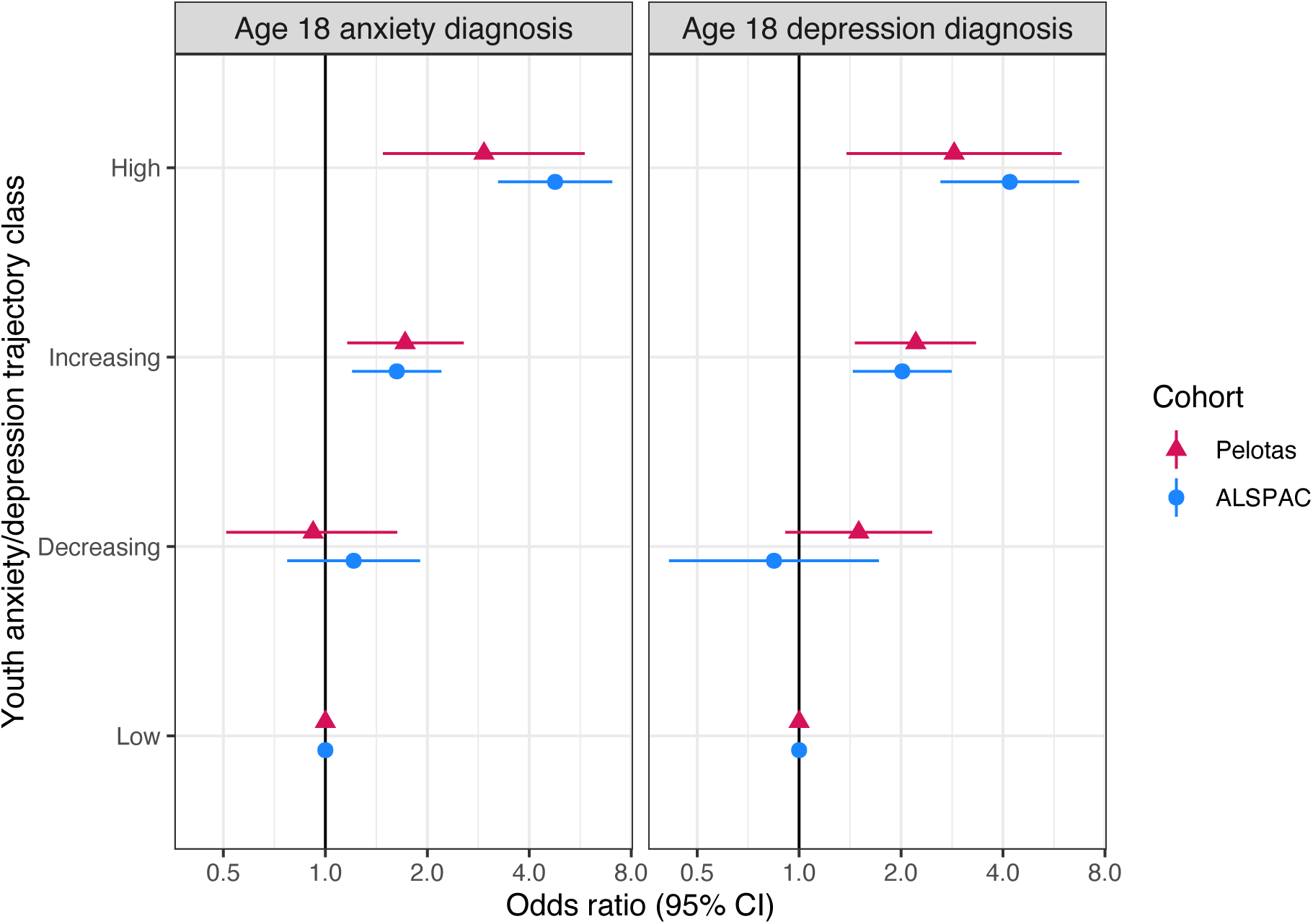
– Results of bias-adjusted three-step association between trajectory class and young adult mental health diagnoses. Note that for simplicity, trajectory class names are shortened. See **Table S8** for full results of statistical tests.

### Secondary analyses

Co-developmental trajectories appear to differ by sex in both cohorts (see **Supplement**).

## Discussion

In this study, we examined the development of anxiety and depression from early childhood to adolescence in two large, population-based birth cohorts from contrasting global contexts: the UK-based ALSPAC cohort and Brazil-based Pelotas 2004 cohort. By combining latent growth models and latent class growth analyses, we sought to characterize the co-development of anxiety conditions and depression, examine whether developmental patterns were comparable across contexts, and test whether trajectories predicted anxiety and depression diagnoses in young adulthood. Across analyses, several findings emerged.

First, despite contextual differences including increased childhood adversity^19^ and poorer access to mental health services in Brazil, we found broad cross-cohort similarities in the prevalence, correlations, and overall developmental patterns of anxiety and depression. In both cohorts, anxiety and depression were moderately correlated at each developmental stage, reflecting shared underlying liability throughout childhood and adolescence.

Despite this, important cross-cohort differences also emerged. In ALSPAC, on average, both anxiety and depression increased linearly from early childhood to adolescence, whereas in Pelotas, anxiety increased steeply but there was not strong evidence for the same increase in depression. Similarly, the development of anxiety and depression were strongly related in ALSPAC but not in Pelotas. Other longitudinal studies from HICs (UK and Portugal) have similarly indicated anxiety and depression co-development across similar developmental periods^37,38^. This observed difference suggests that the Brazilian context may be more influential on increases in anxiety, whereas in the UK, depression and anxiety increase together more gradually. Another possibility is that there are cultural differences in the reporting of anxiety and depression, whereby increases in depression are better recognized and reported in the UK, but less so in Brazil.

The consistent patterns of heterogeneity in the trajectories of anxiety and depression across two socioeconomically and geographically distinct populations suggests a similar set of developmental pathways between contexts. The observed patterns of development, including both increasing and decreasing classes, align with previous literature on developmental trajectories of internalizing problems assessed using questionnaires^39–42^. The increasing class reflects youth whose likelihood of diagnosis rises across the transition from early childhood to adolescence. The decreasing class captures children with an elevated risk of diagnosis in early childhood that attenuates as they mature. Nevertheless, because anxiety and depression levels generally remained low to moderate in the decreasing group in both cohorts, this group does not appear to represent recovery from clinical condition, but rather small shifts in symptom levels within a broadly normal developmental range.

Importantly, and consistent with previous evidence from HICs^39,40^, our findings reveal that childhood trajectories are strong predictors of young adult depression and anxiety diagnoses, with consistent patterns in both cohorts. Young people in both the increasing and high classes had elevated odds of both depression and anxiety at age 18, relative to those in the stable-low class. In contrast, individuals in the decreasing class were not at increased risk for diagnoses in either cohort, suggesting that an elevated likelihood of diagnosis in early childhood may not confer long-term risk if improvement occurs by adolescence. However, this should not overshadow the potential importance of identifying those with symptoms of anxiety in early childhood, as those with the highest risk of young adult diagnoses were already showing elevated levels in early childhood. Furthermore, we were unable to tell whether access to support differed between those whose early childhood symptoms persisted vs. remitted. These findings underscore the prognostic value of longitudinal monitoring^43^ and the importance of early intervention, supported by evidence that treating childhood anxiety reduces adult mental health risk^44,45^.

Together, these findings suggest that despite differences in socioeconomic conditions and access to support, the developmental pathways of anxiety/depression appear broadly comparable across contexts. However, cross-cohort differences in average anxiety and depression change, particularly the stronger increase in anxiety and the weaker anxiety and depression coupling in Pelotas, highlight the need to understand culturally and environmentally specific influences. Second, this study underscores the value of LMIC cohorts for global mental health research. Much of the existing literature is derived from HICs, and our findings show that incorporating LMIC data enables both the validation of global patterns and identification of context-specific aspects. A 2018 systematic review of child and adolescent depression trajectories identified 20 studies, none of which were from LMICs^46^. Expanding such comparative work is essential for building equitable and globally relevant mental health evidence and culturally appropriate treatments.

Several additional strengths should be highlighted. First, identical measures between cohorts allow for direct comparisons. Second, longitudinal birth cohorts allow prospective assessments of anxiety and depression from early childhood to adolescence, a crucial and under-researched developmental period. Many previous studies of anxiety/depression development begin in adolescence, missing the typical onset period for certain anxiety conditions.

Several limitations also warrant consideration. Although both studies used the DAWBA, we cannot rule out idiosyncratic differences in assessment methods between cohorts, including cultural interpretations of symptoms, and parental reporting which could contribute to discrepancies between cohorts. Further, while DAWBA diagnoses have been validated in both contexts, the DAWBA bands have only been validated in HICs. Additionally, the exact assessment ages differed between cohorts, as did the recruitment years (ALSPAC in 1991-2, and Pelotas in 2004). This generational difference may account for some observed differences, reflecting secular trends in youth mental health^47^ rather than contextual factors.

As parent-reported data were available at only three timepoints, we could not model non-linear trajectories, which previous studies indicate may best characterize the development of youth emotional problems^48^. Missing data, while addressed through FIML and MICE, may still introduce bias if unmeasured predictors of attrition exist. Entropy values were low for all models, indicating classification uncertainty, although this was accounted for in class-based associations. Small sample sizes, particularly in Pelotas, prevented modelling of specific anxiety conditions, which each follow distinct developmental courses^49^.

In conclusion, anxiety and depression show broadly similar heterogeneous developmental patterns from childhood to adolescence in UK and Brazilian youth, with trajectories strongly associated with young adult diagnoses. In both cohorts, the persistence versus resolution of early childhood anxiety was critical for later mental health, highlighting the importance of early support and intervention. Our findings suggest shared and potentially context-specific patterns in the development of anxiety and depression, and highlight the importance of tracking change over time. Understanding these developmental pathways is crucial for designing early identification and prevention strategies that can be effective across diverse global contexts.

## Data availability

The informed consent obtained from ALSPAC (Avon Longitudinal Study of Parents and Children) participants does not allow the data to be made available through any third party maintained public repository. Supporting data are available from ALSPAC on request under the approved proposal number, B4518. Full instructions for applying for data access can be found here: http://www.bristol.ac.uk/alspac/researchers/access/. The ALSPAC study website contains details of all available data (http://www.bristol.ac.uk/alspac/researchers/our-data/).

Applications to use Pelotas 2004 data can be made by contacting the researchers of the 2004 Cohort.

## Acknowledgements

This research was funded in whole, or in part, by the Wellcome Trust [309183/Z/24/Z]. This article is based on data from the study “Pelotas Birth Cohort, 2004” conducted by Postgraduate Program in Epidemiology at Universidade Federal de Pelotas, with the collaboration of the Brazilian Public Health Association (ABRASCO). From 2009 to 2013, the Wellcome Trust supported the 2004 birth cohort study. The World Health Organization, National Support Program for Centers of Excellence (PRONEX), Brazilian National Research Council (CNPq), Brazilian Ministry of Health, and Children’s Pastorate supported previous phases of the study. The 2004 Pelotas 11-year follow-up was supported by the Department of Science and Technology (DECIT) of the Brazilian Ministry of Health, CNPq, and the Research Support Foundation of the State of São Paulo (FAPESP; grant number 2014/13864–6). The 15-year follow-up was supported by the DECIT, CNPq, FAPESP (grant number 2020/07730–8), the Research Support Foundation of the State of Rio Grande do Sul (FAPERGS), and the L’Oréal-Unesco-ABC Program for Women in Science in Brazil-2020. The 18-year follow-up was supported by DECIT, CNPq (grant number 409224/2021–9), FAPERGS, L’Oréal-Unesco-ABC Program for Women in Science in Brazil-2020, and the All for Health Institute, São Paulo, Brazil. CNPq supports LTR (308319/2021–4), ISS (303042/2018–4), AJDB, and AM (312746/2021–0). AM also received funding from ESRC and FAPESP (grant number 2023/12905–0). The Wellcome Trust supports MXC (225019/Z/22/Z). HJ is supported by the National Institute for Health and Care Research (NIHR) Bristol Biomedical Research Centre (grant no: NIHR 203315). The views expressed are those of the authors and not necessarily those of the NIHR or the Department of Health and Social Care.

We are extremely grateful to all the families who took part in the ALSPAC study, the midwives for their help in recruiting them, and the whole ALSPAC team, which includes data collection staff, data and administrations staff, technical managers and the technical staff with the Bristol Bioresource Laboratory, based within the University of Bristol. The UK Medical Research Council and Wellcome (Grant ref: MR/Z505924/1) and the University of Bristol provide core support for ALSPAC. This publication is the work of the authors and AS will serve as guarantors for the contents of this paper. A comprehensive list of grants funding is available on the ALSPAC website (http://www.bristol.ac.uk/alspac/external/documents/grant-acknowledgements.pdf). For the purpose of Open Access, the author has applied a CC BY public copyright licence to any Author Accepted Manuscript version arising from this submission.

