## Supplement for "Co-development of anxiety and depression in UK and Brazil youth; a cross-country comparison"

### Supplementary Material

**Contents**

1. Supplementary Text
2. Supplementary Tables
3. Supplementary Figures
4. Deviations from pre-registration
5. Analysis Scripts

#### Supplementary Text

##### Ethics

Ethical approval for the study was obtained from the ALSPAC Ethics and Law Committee and the Local Research Ethics Committees. Informed consent for the use of all data collected was obtained from participants following the recommendations of the ALSPAC Ethics and Law Committee at the time. Participants can contact the study team at any time to retrospectively withdraw consent for their data to be used. Study participation is voluntary and during all data collection sweeps, information was provided on the intended use of data.

The 2004 Pelotas Birth Cohort follow-ups were approved by the Federal University of Pelotas Medical School Research Ethics Committee.

##### Growth curve modelling methods

Within-wave residual correlations between anxiety and depression were included to account for shared rater and occasion-specific variance. These correlations were present in both cohorts, but stronger in Pelotas than in ALSPAC, indicating that occasion-specific effects contributed to the observed relationship between anxiety and depression. Correlations between latent variables (intercepts and slopes) were included based on modification indices and best model fit criteria (assessed using comparative fit index (CFI), Root Mean Square Error of Approximation (RMSEA) and Standardized Root Mean Square Residual (SRMR)). In ALSPAC, this resulted in all slope/intercept correlations being included, and in Pelotas all correlations, except between depression slope and anxiety and depression intercepts, were included. Due to skewed ordinal data, models were fitted using Weighted Least Squares Mean and Variance adjusted (WLSMV) and multiple imputation was performed to handle missing data.

##### Growth mixture modelling methods

Starting with a single k-class model, k+1 solutions were fitted until the optimal model was reached, as determined primarily using Lo-Mendell-Rubin (LMR) tests, supported by Akaike information criterion (AIC), Bayesian information criterion (BIC), smallest class size information, as well as the plausibility and interpretability of classes. The LMR test quantifies whether a k-class model substantially improves model fit enough to justify the additional parameters introduced compared to the k-1 class model. Lower values on AIC and BIC indicate better model fit. Once the optimal model was determined, we repeated with an increased number of random starts (n=500), to ensure no problems with the local maxima. LCGA models were fitted using maximum likelihood estimator and used full information maximum likelihood (FIML) to handle missing data (different to LGM as mixture modelling is not compatible with WLSMV estimator).

##### Missing data and multiple imputation procedure

This imputed sample includes individuals for whom anxiety or depression data were available from at least one timepoint (N=9,586 in ALSPAC and N=3,815 in Pelotas). The multiple imputation model included all variables (predictors and outcomes) in the analyses, as well as known predictors of missingness. In ALSPAC this included: maternal age at birth, maternal education, parity, smoking during pregnancy, and home ownership status. In Pelotas, this included: maternal age, maternal marital status, maternal education, family income, smoking during pregnancy, drinking alcohol during pregnancy, offspring birthweight and maternal skin colour. For each cohort, 200 datasets were imputed in R using the package ‘mice’ and the predictive mean matching method. **ETable 1** and **2** report the proportion of missing data for each variable in both cohorts.

Anxiety-only and depression-only trajectory models

For depression, the 3-class solution was optimal in both ALSPAC and Pelotas 2004. Both cohorts showed a stable low depression class (ALSPAC = 55%, Pelotas = 37%), an intermediate class (ALSPAC = 43%, Pelotas = 59%, although this class was slightly increasing in ALSPAC and stable in Pelotas) and a high-increasing class (ALSPAC = 2%, Pelotas = 4%). For model fit and plots see **eTable 6-7** and **eFigure 2**.

For anxiety, in ALSPAC the 4-class model was the best-fitting solution, compared to the 3-class model in Pelotas 2004. Both cohorts showed a stable low likelihood of anxiety diagnosis class (ALSPAC = 40%, Pelotas = 58%) and an intermediate-increasing class (ALSPAC = 13%; Pelotas = 39%). ALSPAC also showed a stable high (4%) and a large intermediate-decreasing class (43%), whereas Pelotas only showed a high-decreasing class (3%). For model fit and plots see **eTable 8-9** and **eFigure 3**.

Prevalence of anxiety conditions in trajectory classes

The prevalence of each anxiety condition at early childhood, middle childhood and adolescence in each of the anxiety/depression trajectory groups, in both cohorts is presented in **eFigure 4.** In both cohorts, as expected, the prevalence of each anxiety condition was highest in the high class and lowest in the low class across all ages. The decreasing class showed the second-highest prevalence for most anxiety conditions in early and middle childhood (with the exception of GAD), whereas during adolescence the increasing class exhibited the second-highest prevalence across all anxiety conditions.

Sex-stratified anxiety and depression trajectories

Sex-stratified descriptive statistics are presented in **eTable 14**. Females in Pelotas showed higher rates of any anxiety condition at all timepoints, particularly specific phobias, compared to ALSPAC. For males, there were higher rates of depression in ALSPAC and higher rates of specific phobias in Pelotas.

Model fit statistics for parallel-process LCGA of anxiety and depression in males and females in both cohorts are presented in the **eTables 15-18** and trajectory figures in **eFigure 5-6.** In males, the 4-class solution was the best fit in ALSPAC and the 3-class in Pelotas. In both cohorts there was a majority class with stable-low anxiety and depression (ALSPAC 46%; Pelotas 55%), and a small class with stable high depression but, in ALSPAC also with stable high anxiety in this group, compared to decreasing anxiety in Pelotas (ALSPAC 4%; Pelotas 3%). In ALSPAC, there was a class with intermediate-decreasing anxiety and depression (30%), and a class with intermediate-increasing depression and stable anxiety (21%). In Pelotas the final class showed stable intermediate anxiety and depression across development (42%). Interestingly, in both cohorts there was no class showing rapidly increasing anxiety or depression, as in females, detailed below.

In females, both the 4 and 5-class solutions showed good fit in ALSPAC but upon visualising trajectories, the 4-class solution was deemed optimal due to two very similar moderate groups in the 5-class solution. In Pelotas, the 4-class solution was also the best fit. Classes followed a similar pattern in both cohorts, although the proportion of sample in each class varied – i) a stable-low class (ALSPAC 33%; Pelotas 48%), ii) an intermediate-increasing class (ALSPAC 13%; Pelotas 31%), iii) a high-increasing class (ALSPAC 11%; Pelotas 4%), and finally, iv) a class showing intermediate-decreasing anxiety and stable-low depression in ALSPAC (43%) and a similar class showing intermediate-decreasing anxiety and depression in Pelotas (18%).

Key differences in trajectories between males and females in ALSPAC were that there was no class showing increasing anxiety in males, but there were two classes characterised by this in females. Similarly, in Pelotas there was no class characterised by increasing anxiety or increasing depression in males, but two classes characterised by this in females.

Unimputed (available cases)

Unimputed (available case) descriptive statistics with can be found in **eTable 19**, showing a similar pattern of results as in the imputed data.

#### Supplementary Tables

**eTable 1**. N and proportion of missing data in ALSPAC (Total N = 9,586)

| Variable | N complete | N Missing | % Missing |
|---|---|---|---|
| Sex | 9562 | 24 | 0.3% |
| Maternal age | 9205 | 381 | 4.0% |
| Smoking during pregnancy | 9020 | 566 | 5.9% |
| Parity | 8893 | 693 | 7.2% |
| Maternal education | 8874 | 712 | 7.4% |
| Home ownership | 8670 | 916 | 9.6% |
| Any anxiety DAWBA band age 7 | 8138 | 1448 | 15.1% |
| Phobia DAWBA band age 7 | 8123 | 1463 | 15.3% |
| Social anxiety DAWBA band age 7 | 8110 | 1476 | 15.4% |
| Separation anxiety DAWBA band age 7 | 8102 | 1484 | 15.5% |
| GAD DAWBA band age 7 | 8090 | 1496 | 15.6% |
| Depression DAWBA band age 7 | 7980 | 1606 | 16.8% |
| Any anxiety DAWBA band age 10 | 7717 | 1869 | 19.5% |
| Social anxiety DAWBA band age 10 | 7691 | 1895 | 19.8% |
| Phobia DAWBA band age 10 | 7682 | 1904 | 19.9% |
| GAD DAWBA band age 10 | 7667 | 1919 | 20.0% |
| Depression DAWBA band age 10 | 7553 | 2033 | 21.2% |
| Separation anxiety DAWBA band age 10 | 7424 | 2162 | 22.6% |
| Phobia DAWBA band age 13 | 7010 | 2576 | 26.9% |
| Any anxiety DAWBA band age 13 | 7010 | 2576 | 26.9% |
| Social anxiety DAWBA band age 13 | 6981 | 2605 | 27.2% |
| GAD DAWBA band age 13 | 6964 | 2622 | 27.4% |
| Depression DAWBA band age 13 | 6866 | 2720 | 28.4% |
| Separation anxiety DAWBA band age 13 | 6430 | 3156 | 32.9% |
| CIS-R diagnoses age 18 | 4145 | 5441 | 56.8% |

**eTable 2**. N and proportion of missing data in Pelotas 2004 (total N =3,815)

| Variable | Complete N | Missing N | Missing % |
|---|---|---|---|
| Sex | 3815 | 0 | 0 |
| Smoking during pregnancy | 3814 | 1 | 0.03% |
| Marital status | 3814 | 1 | 0.03% |
| Family income | 3814 | 1 | 0.03% |
| Alcohol during pregnancy | 3814 | 1 | 0.03% |
| Maternal ethnicity | 3814 | 1 | 0.03% |
| Maternal age | 3812 | 3 | 0.08% |
| Maternal education | 3779 | 36 | 0.94% |
| Any anxiety DAWBA band age 6 | 3584 | 231 | 6.1% |
| Depression DAWBA band age 6 | 3584 | 231 | 6.1% |
| Social anxiety DAWBA band age 6 | 3584 | 231 | 6.1% |
| Separation anxiety DAWBA band age 6 | 3583 | 232 | 6.1% |
| Phobia DAWBA band age 6 | 3582 | 233 | 6.1% |
| GAD DAWBA band age 6 | 3582 | 233 | 6.1% |
| Any anxiety DAWBA band age 11 | 3563 | 252 | 6.6% |
| Depression DAWBA band age 11 | 3563 | 252 | 6.6% |
| Phobia DAWBA band age 11 | 3563 | 252 | 6.6% |
| Social anxiety DAWBA band age 11 | 3563 | 252 | 6.6% |
| GAD DAWBA band age 11 | 3563 | 252 | 6.6% |
| Separation anxiety DAWBA band age 11 | 3562 | 253 | 6.6% |
| Any anxiety DAWBA band age 15 | 1941 | 1874 | 49.1% |
| Depression DAWBA band age 15 | 1941 | 1874 | 49.1% |
| Phobia DAWBA band age 15 | 1941 | 1874 | 49.1% |
| GAD DAWBA band age 15 | 1941 | 1874 | 49.1% |
| Social anxiety DAWBA band age 15 | 1941 | 1874 | 49.1% |
| Separation anxiety DAWBA band age 15 | 1936 | 1879 | 49.1% |
| Any anxiety MINI age 18 | 3166 | 649 | 17.0% |
| Depression MINI age 18 | 3168 | 647 | 17.0% |

**eTable 3 –** Anxiety and depression prevalence rates in young adulthood (age 18) in ALSPAC and Pelotas cohorts. Diagnoses were derived using the CIS-R in ALSPAC and the MINI in Pelotas 2004. N/A indicates where this was not measured in the sample.

| **Diagnosis** | **ALSPAC (N = 9,586)** | | **Pelotas (N = 3,815)** | |
|---|---|---|---|---|
| | **N** | **%** | **N** | **%** |
| Depression | 834 | 8.7 | 634 | 16.6 |
| Any anxiety condition | 1130 | 11.8 | 647 | 17.8 |
| Specific phobia | 443 | 4.6 | N/A | N/A |
| Social anxiety | 215 | 2.2 | N/A | N/A |
| GAD | 621 | 6.5 | N/A | N/A |
| Anxiety depression comorbidity | 409 | 4.3 | 302 | 7.9 |

**eTable 4** – Frequency distribution of DAWBA band variables in ALSPAC and Pelotas. Note that the 3% band is not included for depression.

| DAWBA band | <0.1% | | 0.50% | | 3% | | 15% | | 50% | | >70% | |
|---|---|---|---|---|---|---|---|---|---|---|---|---|
| | N | % | N | % | N | % | N | % | N | % | N | % |
| **ALSPAC (N = 9,586)** | | | | | | | | | | | | |
| **anyanx7** | ^ | | 4193 | 43.7% | 4671 | 48.7% | 552 | 5.8% | 94 | 1.0% | 76 | 0.8% |
| **anyanx10** | ^ | | 3545 | 37.0% | 5174 | 54.0% | 648 | 6.8% | 123 | 1.3% | 96 | 1.0% |
| **anyanx13** | 18 | 0.2% | 3981 | 41.5% | 4877 | 50.9% | 525 | 5.5% | 120 | 1.3% | 64 | 0.7% |
| **dep7** | 5917 | 61.7% | 3376 | 35.2% | *NA* | | 231 | 2.4% | 62 | 0.6% | ^ | |
| **dep10** | 5624 | 58.7% | 3522 | 36.7% | *NA* | | 332 | 3.5% | 99 | 1.0% | 9 | 0.1% |
| **dep13** | 5489 | 57.3% | 3516 | 36.7% | *NA* | | 466 | 4.9% | 90 | 0.9% | 25 | 0.3% |
| **Pelotas (N = 3,815)** | | | | | | | | | | | | |
| **anyanx6** | ^ | | 3014 | 79.0% | 545 | 14.3% | 175 | 4.6% | 46 | 1.2% | 35 | 0.9% |
| **anyanx11** | ^ | | 2792 | 73.2% | 664 | 17.4% | 236 | 6.2% | 90 | 2.4% | 32 | 0.8% |
| **anyanx15** | ^ | | 2726 | 71.5% | 858 | 22.5% | 165 | 4.3% | 56 | 1.5% | 10 | 0.3% |
| **dep6** | 3022 | 79.2% | 685 | 17.9% | *NA* | | 92 | 2.4% | 16 | 0.4% | ^ | |
| **dep11** | 2970 | 77.9% | 760 | 19.9% | *NA* | | 70 | 1.8% | 15 | 0.4% | ^ | |
| **dep15** | 2817 | 73.8% | 808 | 21.2% | *NA* | | 136 | 3.6% | 42 | 1.1% | 12 | 0.3% |

*^Due to disclosure protocols, bands with N<5 are not given as exact values, with Ns collapsed across bands.*

**eTable 5 -** Results of growth curve modelling in both cohorts. Intercept means are not presented due to being fixed at zero for model specification. Fully standardized estimates (STDYX) are presented alongside unstandardized estimates, to facilitate comparison between cohorts.

| Parameter | ALSPAC | | | | Pelotas | | | |
|---|---|---|---|---|---|---|---|---|
| | **Un-standardized estimate** | **SE** | **P-value** | **Fully standardised estimate** | **Un-standardized estimate** | **SE** | **P-value** | **Fully standardised estimate** |
| **Slope mean** | | | | | | | | |
| S_DEP | 0.016 | 0.004 | <0.001 | 0.17 | 0.009 | 0.006 | 0.13 | 0.80 |
| S_ANX | 0.005 | 0.003 | 0.048 | 0.05 | 0.042 | 0.005 | <0.001 | 0.49 |
| **Intercept variances** | | | | | | | | |
| I_DEP | 0.423 | 0.037 | <0.001 | *NA* | 0.311 | 0.026 | <0.001 | *NA* |
| I_ANX | 0.641 | 0.028 | <0.001 | *NA* | 0.624 | 0.060 | <0.001 | *NA* |
| **Slope variances** | | | | | | | | |
| S_DEP | 0.009 | 0.002 | <0.001 | *NA* | <0.001 | 0.001 | 0.84 | *NA* |
| S_ANX | 0.011 | 0.002 | <0.001 | *NA* | 0.008 | 0.001 | <0.001 | *NA* |
| **Intercept–Intercept correlation** | | | | | | | | |
| I_DEP ↔ I_ANX | 0.364 | 0.023 | <0.001 | 0.70 | 0.235 | 0.025 | <0.001 | 0.52 |
| **Slope–Slope correlation** | | | | | | | | |
| S_ANX ↔ S_DEP | 0.007 | 0.001 | <0.001 | 0.72 | 0.001 | 0.001 | 0.31 | 0.88 |
| **Within-construct intercept–slope correlations** | | | | | | | | |
| I_DEP ↔ S_DEP | −0.009 | 0.008 | 0.25 | -0.14 | *NA* | *NA* | *NA* | *NA* |
| I_ANX ↔ S_ANX | −0.037 | 0.006 | <0.001 | -0.45 | -0.052 | 0.008 | <0.001 | -0.75 |
| **Cross-domain intercept–slope correlations** | | | | | | | | |
| I_DEP ↔ S_ANX | −0.020 | 0.005 | <0.001 | -0.30 | -0.012 | 0.004 | 0.003 | -0.25 |
| I_ANX ↔ S_DEP | −0.017 | 0.005 | <0.001 | -0.23 | *NA* | *NA* | *NA* | *NA* |
| **Within-wave residual correlations** | | | | | | | | |
| DEP7 ↔ ANX7 | 0.063 | 0.025 | 0.009 | 0.15 | 0.264 | 0.031 | <0.001 | 0.52 |
| DEP10 ↔ ANX10 | 0.158 | 0.015 | <0.001 | 0.26 | 0.191 | 0.027 | <0.001 | 0.31 |
| DEP13 ↔ ANX13 | 0.084 | 0.028 | 0.002 | 0.18 | 0.218 | 0.047 | <0.001 | 0.45 |

*ALSPAC model fit: Comparative fit index (CFI) = 0.99, Root Mean Square Error of Approximation (RMSEA) = 0.038, Standardized Root Mean Square Residual (SRMR) = 0.011*
*Pelotas model fit: CFI = 0.99, RMSEA =0.021, SRMR = 0.015*

**eTable 6** – Model fit of depression LCGA in ALSPAC

| **Number of classes** | **1** | **2** | **3** | **4** |
|---|---|---|---|---|
| **Parameters** | 5 | 8 | 11 | 14 |
| **LL** | -18659 | -18097 | -18050 | -18045 |
| **AIC** | 37328 | 36210 | 36122 | 36119 |
| **BIC** | 37364 | 36267 | 36201 | 36219 |
| **Entropy** | - | 0.43 | 0.56 | 0.65 |
| **ALMR p-value** | - | <0.0001 | <0.0001 | 0.04 |
| **BLRT p-value** | - | <0.0001 | <0.0001 | 0.05 |
| **Class 1 N (%)** | 9509 (100%) | 3426 (36%) | 4104 (43%) | 188 (2%) |
| **Class 2 N (%)** | | 6083 (64%) | 5207 (55%) | 5226 (55%) |
| **Class 3 N (%)** | | | 198 (2%) | 4082 (43%) |
| **Class 4 N (%)** | | | | 13 (0.1%) |

**eTable 7** – Model fit of depression LCGA in Pelotas 2004

| **Number of classes** | **1** | **2** | **3** | **4** |
|---|---|---|---|---|
| **Parameters** | 5 | 8 | 11 | 14 |
| **LL** | -5800 | -5686 | -5679 | -5674 |
| **AIC** | 11609 | 11389 | 11380 | 11375 |
| **BIC** | 11640 | 11439 | 11448 | 11462 |
| **Entropy** | - | 0.51 | 0.49 | 0.57 |
| **ALMR p-value** | - | <0.001 | 0.038 | 0.24 |
| **BLRT p-value** | - | <0.001 | <0.001 | 0.04 |
| **Class 1 N (%)** | 3816 (100%) | 801 (21%) | 2245 (59%) | 39 (1%) |
| **Class 2 N (%)** | | 3014 (79%) | 156 (4%) | 134 (4%) |
| **Class 3 N (%)** | | | 1414 (37%) | 1590 (42%) |
| **Class 4 N (%)** | | | | 2053 (54%) |

**eTable 8** – Model fit of anxiety LCGA in ALSPAC

| **Number of classes** | **1** | **2** | **3** | **4** | **5** |
|---|---|---|---|---|---|
| **Parameters** | 6 | 9 | 12 | 15 | 18 |
| **LL** | -21758 | -20742 | -20558 | -20537 | -20529 |
| **AIC** | 43527 | 41501 | 41141 | 41105 | 41094 |
| **BIC** | 43570 | 41566 | 41227 | 41212 | 41223 |
| **Entropy** | - | 0.49 | 0.62 | 0.53 | 0.59 |
| **ALMR p-value** | - | <0.0001 | <0.0001 | <0.0001 | <0.0001 |
| **BLRT p-value** | - | <0.0001 | <0.0001 | <0.0001 | <0.0001 |
| **Class 1 N (%)** | 9586 (100%) | 4762 (50%) | 3829 (40%) | 3802 (40%) | 4082 (43%) |
| **Class 2 N (%)** | | 4824 (50%) | 5325 (56%) | 1264 (13%) | 3817 (40%) |
| **Class 3 N (%)** | | | 432 (5%) | 431 (4%) | 1237 (13%) |
| **Class 4 N (%)** | | | | 4090 (43%) | 17 (0.2%) |
| **Class 5 N (%)** | | | | | 432 (5%) |

**eTable 9** – Model fit of anxiety LCGA in Pelotas 2004

| **Number of classes** | **1** | **2** | **3** | **4** |
|---|---|---|---|---|
| **Parameters** | 5 | 8 | 11 | *Error** |
| **LL** | -7020 | -6866 | -6859 | |
| **AIC** | 14050 | 13748 | 13740 | |
| **BIC** | 14081 | 13798 | 13808 | |
| **Entropy** | - | 0.53 | 0.49 | |
| **ALMR p-value** | - | <0.001 | 0.0322 | |
| **BLRT p-value** | - | <0.001 | <0.001 | |
| **Class 1 N (%)** | 3816 (100%) | 843 (22%) | 1471 (39%) | |
| **Class 2 N (%)** | | 2972 (78%) | 2216 (58%) | |
| **Class 3 N (%)** | | | 128 (3%) | |

**Unable to run 4-class model due to error*

**eTable 10 –** Model fit statistics for the parallel-process LCGA (1–5 classes) of anxiety and depression in the Pelotas 2004 cohort. The 4-class solution showed optimal fit.

| **Number of Classes** | **1** | **2** | **3** | **4** | **5** |
|---|---|---|---|---|---|
| **Parameters** | 10 | 15 | 20 | **25** | 30 |
| **LL** | -12820 | -12315 | -12266 | **-12223** | -12209 |
| **AIC** | 25659 | 24661 | 24573 | **24495** | 24478 |
| **BIC** | 25722 | 24755 | 24698 | **24652** | 24665 |
| **Entropy** | - | 0.54 | 0.48 | **0.54** | 0.6 |
| **ALMR p-value** | - | <0.001 | 0.0039 | **0.011** | 0.26 |
| **BLRT p-value** | - | <0.001 | <0.001 | **<0.001** | <0.001 |
| **Class 1 N (%)** | 3815 (100%) | 2573 (67%) | 941 (25%) | **1073 (28%)** | 487 (13%) |
| **Class 2 N (%)** | | 1242 (33%) | 2225 (58%) | **576 (15%)** | 1038 (27%) |
| **Class 3 N (%)** | | | 649 (17%) | **2056 (54%)** | 2090 (55%) |
| **Class 4 N (%)** | | | | **110 (3%)** | 99 (3%) |
| **Class 5 N (%)** | | | | | 102 (3%) |

*Log-likelihood (LL), Akaike information criterion (AIC), Bayesian information criterion (BIC), Adjusted Lo-Mendell-Rubin (ALMR), Bootstrap Likelihood Ratio Test (BLRT)*

**eTable 11** - Model fit statistics for the parallel-process LCGA (1–7 classes) of anxiety and depression in the ALSPAC cohort. For 2-6 class models, the ALMR p-value was ≤0.001 and smallest class size >1%, however visualisation of classes determined the 4-class solution to be optimal.

| **Number of Classes** | **1** | **2** | **3** | **4** | **5** | **6** | **7** |
|---|---|---|---|---|---|---|---|
| **Parameters** | 11 | 16 | 21 | **26** | 31 | 36 | 41 |
| **LL** | -40417 | -38239 | -37901 | **-37794** | -37719 | -37664 | -37621 |
| **AIC** | 80855 | 76510 | 75843 | **75639** | 75500 | 75399 | 75324 |
| **BIC** | 80934 | 76625 | 75994 | **75826** | 75722 | 75658 | 75618 |
| **Entropy** | - | 0.57 | 0.64 | **0.55** | 0.48 | 0.48 | 0.51 |
| **ALMR p-value** | - | <0.0001 | <0.0001 | **<0.0001** | 0.0063 | 0.0005 | 0.001 |
| **BLRT p-value** | - | <0.0001 | <0.0001 | **<0.0001** | <0.0001 | <0.0001 | <0.0001 |
| **Class 1 N (%)** | 9586 (100%) | 5251 (55%) | 5002 (52%) | **3967 (41%)** | 2746 (29%) | 2495 (26%) | 2480 (26%) |
| **Class 2 N (%)** | | 4335 (45%) | 3967 (41%) | **2988 (31%)** | 2331 (24%) | 2146 (22%) | 2291 (24%) |
| **Class 3 N (%)** | | | 617 (6%) | **2158 (23%)** | 2186 (23%) | 2054 (21%) | 1832 (19%) |
| **Class 4 N (%)** | | | | **473 (5%)** | 2061 (22%) | 1799 (19%) | 1751 (18%) |
| **Class 5 N (%)** | | | | | 261 (3%) | 839 (9%) | 881 (9%) |
| **Class 6 N (%)** | | | | | | 253 (3%) | 228 (2%) |
| **Class 7 N (%)** | | | | | | | 123 (1%) |

*Log-likelihood (LL), Akaike information criterion (AIC), Bayesian information criterion (BIC), Adjusted Lo-Mendell-Rubin (ALMR), Bootstrap Likelihood Ratio Test (BLRT)*

**eTable 12 –** Slope estimates from parallel-process latent class growth models of anxiety and depression in ALSPAC and Pelotas 2004 cohorts.

| Cohort | Class | Anxiety Slope | | | Depression Slope | | |
|---|---|---|---|---|---|---|---|
| | | Estimate | SE | P-value | Estimate | SE | P-value |
| ALSPAC | Stable-low | 0.063 | 0.014 | <0.001 | 0.077 | 0.015 | <0.001 |
| | Intermediate-increasing | 0.114 | 0.028 | <0.001 | 0.152 | 0.025 | <0.001 |
| | Intermediate-decreasing | -0.19 | 0.033 | <0.001 | -0.166 | 0.029 | <0.001 |
| | Persistent-high anxiety/increasing depression | -0.042 | 0.037 | 0.26 | 0.104 | 0.046 | 0.024 |
| Pelotas | Stable-low | -0.037 | 0.034 | 0.27 | -0.009 | 0.04 | 0.82 |
| | Intermediate-increasing | 0.187 | 0.031 | <0.001 | 0.189 | 0.039 | <0.001 |
| | Intermediate-decreasing | -0.154 | 0.049 | 0.002 | -0.196 | 0.06 | 0.001 |
| | Persistent-high | -0.004 | 0.07 | 0.95 | 0.129 | 0.11 | 0.24 |

**eTable 13** – Results of bias adjusted 3-step association between trajectory class and young adult mental health diagnoses. Note, the persistent-high anxiety/increasing depression class in ALSPAC and the persistent-high anxiety and depression class in Pelotas are both referred to as ‘high’.

| Age 18 diagnosis | Cohort | Stable low class | Intermediate-decreasing class | | | Intermediate-increasing class | | | High class | | |
|---|---|---|---|---|---|---|---|---|---|---|---|
| | | OR | OR | 95% CI | P-value | OR | 95% CI | P-value | OR | 95% CI | P-value |
| Anxiety | ALSPAC | 1.00 (ref) | 1.21 | 0.77, 1.90 | 0.40 | 1.63 | 1.20, 2.20 | 0.002 | 4.77 | 3.24, 7.03 | 0.002 |
| | Pelotas | 1.00 (ref) | 0.92 | 0.51, 1.63 | 0.77 | 1.72 | 1.16, 2.56 | 0.008 | 2.94 | 1.48, 5.83 | 0.002 |
| Depression | ALSPAC | 1.00 (ref) | 0.84 | 0.41, 1.72 | 0.64 | 2.02 | 1.44, 2.82 | <0.001 | 4.19 | 2.61, 6.71 | <0.001 |
| | Pelotas | 1.00 (ref) | 1.5 | 0.91, 2.47 | 0.11 | 2.21 | 1.46, 3.33 | <0.001 | 2.87 | 1.38, 5.97 | 0.005 |

**eTable 14** – Sex-stratified anxiety and depression diagnoses across early childhood, middle childhood, adolescence and young adulthood in ALSPAC and Pelotas 2004 birth cohorts. Diagnoses at early childhood, middle childhood and adolescence were derived from the DAWBA. Diagnoses in young adulthood are derived using the CIS-R in ALSPAC and the MINI in Pelotas 2004. N/A indicates where this was not measured in the sample.

| Diagnosis | Developmental stage | Females | | | | Males | | | |
|---|---|---|---|---|---|---|---|---|---|
| | | ALSPAC (total N = 4702) | | Pelotas (total N =1832) | | ALSPAC (total N = 4860) | | Pelotas (total N = 1983) | |
| | | N | % | N | % | N | % | N | % |
| Depression | Childhood | 27 | 0.6% | 7 | 0.4% | 35 | 0.7% | 11 | 0.5% |
| | Pre-adolescence | 45 | 0.9% | 5 | 0.3% | 64 | 1.3% | 9 | 0.5% |
| | Adolescence | 47 | 1.0% | 38 | 2.1% | 72 | 1.5% | 19 | 1.0% |
| | Young adulthood | 511 | 10.9% | 446 | 24.4% | 321 | 6.6% | 188 | 9.5% |
| Any anxiety disorder | Childhood | 83 | 1.8% | 38 | 2.1% | 106 | 2.2% | 46 | 2.3% |
| | Pre-adolescence | 128 | 2.7% | 68 | 3.7% | 144 | 3.0% | 69 | 3.5% |
| | Adolescence | 107 | 2.3% | 44 | 2.4% | 122 | 2.5% | 31 | 1.6% |
| | Young adulthood | 628 | 13.4% | 461 | 25.2% | 498 | 10.3% | 186 | 9.4% |
| Specific phobia | Childhood | 52 | 1.1% | 23 | 1.2% | 57 | 1.2% | 30 | 1.5% |
| | Pre-adolescence | 52 | 1.1% | 47 | 2.6% | 65 | 1.3% | 48 | 2.4% |
| | Adolescence | 23 | 0.5% | 33 | 1.8% | 34 | 0.7% | 25 | 1.3% |
| | Young adulthood | 246 | 5.2% | N/A | N/A | 196 | 4.0% | N/A | N/A |
| Social phobia | Childhood | 6 | 0.1% | <5 | <0.3% | 5 | 0.1% | <5 | <0.3% |
| | Pre-adolescence | 17 | 0.4% | <5 | <0.3% | 22 | 0.5% | <5 | <0.3% |
| | Adolescence | 24 | 0.5% | 9 | 0.5% | 32 | 0.6% | <5 | <0.3% |
| | Young adulthood | 120 | 2.6% | N/A | N/A | 94 | 1.9% | N/A | N/A |
| GAD | Childhood | 5 | 0.1% | 0 | 0% | 15 | 0.3% | 0 | 0% |
| | Pre-adolescence | 21 | 0.4% | <5 | <0.3% | 37 | 0.8% | <5 | <0.3% |
| | Adolescence | 30 | 0.6% | <5 | <0.3% | 39 | 0.8% | 0 | 0% |
| | Young adulthood | 344 | 7.3% | N/A | N/A | 274 | 5.6% | N/A | N/A |
| Separation anxiety | Childhood | 39 | 0.8% | 17 | 0.9% | 46 | 0.9% | 19 | 1.0% |
| | Pre-adolescence | 60 | 1.3% | 20 | 1.1% | 71 | 1.5% | 22 | 1.1% |
| | Adolescence | 57 | 1.2% | 5 | 0.3% | 58 | 1.2% | <5 | 0.1% |
| | Young adulthood | N/A | N/A | N/A | N/A | N/A | N/A | N/A | N/A |
| Anxiety depression comorbidity | Childhood | 10 | 0.2% | 5 | 0.3% | 12 | 0.2% | 5 | 0.3% |
| | Pre-adolescence | 15 | 0.3% | <5 | <0.3% | 28 | 0.6% | <5 | <0.3% |
| | Adolescence | 20 | 0.4% | 5 | 0.3% | 32 | 0.7% | <5 | <0.3% |
| | Young adulthood | 245 | 5.2% | 233 | 12.7% | 163 | 3.4% | 70 | 3.5% |

**eTable 15** - Model fit of parallel LCGA in ALSPAC – Males only (N=4860)

| **Number of classes** | 1 | 2 | 3 | **4** | 5 |
|---|---|---|---|---|---|
| **Parameters** | 11 | 16 | 21 | **26** | 31 |
| **LL** | -19953 | -18921 | -18738 | **-18689** | -18650 |
| **AIC** | 39928 | 37874 | 37517 | **37429** | 37363 |
| **BIC** | 39999 | 37978 | 37653 | **37598** | 37564 |
| **Entropy** | | 0.57 | 0.66 | **0.56** | 0.61 |
| **ALMR p-value** | | <0.001 | <0.001 | **0.0055** | 0.057 |
| **BLRT p-value** | | <0.001 | <0.001 | **<0.001** | <0.001 |
| **Class 1 N (%)** | 4860 (100%) | 2840 (58%) | 2373 (49%) | **2228 (46%)** | 2010 (41%) |
| **Class 2 N (%)** | | 2020 (42%) | 207 (4%) | **1011 (21%)** | 53 (1%) |
| **Class 3 N (%)** | | | 2280 (47%) | **1449 (30%)** | 1810 (37%) |
| **Class 4 N (%)** | | | | **172 (4%)** | 197 (4%) |
| **Class 5 N (%)** | | | | | 790 (16%) |

**eTable 16** - Model fit of parallel LCGA in Pelotas 2004 – Males only (N=1983)

| **Number of classes** | 1 | 2 | **3** | 4 |
|---|---|---|---|---|
| **Parameters** | 10 | 15 | **20** | 25 |
| **LL** | -6500 | -6219 | **-6188** | -6173 |
| **AIC** | 13019 | 12468 | **12417** | 12395 |
| **BIC** | 13075 | 12552 | **12529** | 12535 |
| **Entropy** | | 0.55 | **0.61** | 0.60 |
| **ALMR p-value** | | <0.001 | **0.04** | 0.09 |
| **BLRT p-value** | | <0.001 | **<0.001** | <0.001 |
| **Class 1 N (%)** | 1983 (100%) | 1339 (68%) | **838 (42%)** | 633 (32%) |
| **Class 2 N (%)** | | 644 (33%) | **1087 (55%)** | 170 (9%) |
| **Class 3 N (%)** | | | **57.8 (3%)** | 1130 (57%) |
| **Class 4 N (%)** | | | | 50 (3%) |

**eTable 17** - Model fit of parallel LCGA in ALSPAC – Females only (N=4702)

| **Number of classes** | 1 | 2 | 3 | **4** | 5 | 6 |
|---|---|---|---|---|---|---|
| **Parameters** | 11 | 16 | 21 | **26** | 31 | 36 |
| **LL** | -20213 | -19118 | -18957 | **-18903** | -18861 | -18826 |
| **AIC** | 40448 | 38268 | 37956 | **37859** | 37784 | 37723 |
| **BIC** | 40519 | 38372 | 38091 | **38026** | 37984 | 37956 |
| **Entropy** | | 0.57 | 0.60 | **0.54** | 0.51 | 0.50 |
| **ALMR p-value** | | <0.001 | <0.001 | **0.033** | 0.015 | 0.18 |
| **BLRT p-value** | | <0.001 | <0.001 | **<0.001** | <0.001 | <0.001 |
| **Class 1 N (%)** | 4702 (100%) | 2376 (51%) | 480 (10%) | **523 (11%)** | 691 (15%) | 971 (21%) |
| **Class 2 N (%)** | | 2326 (49%) | 1634 (35%) | **1573 (33%)** | 353 (8%) | 1237 (26%) |
| **Class 3 N (%)** | | | 2588 (55%) | **594 (13%)** | 1544 (33%) | 1210 (26%) |
| **Class 4 N (%)** | | | | **2012 (43%)** | 1516 (32%) | 700 (15%) |
| **Class 5 N (%)** | | | | | 598 (13%) | 144 (3%) |
| **Class 6 N (%)** | | | | | | 439 (9%) |

**eTable 18** - Model fit of parallel LCGA in Pelotas 2004 – Females only (N=1832)

| **Number of classes** | 1 | 2 | 3 | **4** | 5 |
|---|---|---|---|---|---|
| **Parameters** | 10 | 15 | 20 | **25** | 30 |
| **LL** | -6309 | -6083 | -6051 | **-6025** | -6014 |
| **AIC** | 12638 | 12195 | 12141 | **12100** | 12088 |
| **BIC** | 12693 | 12278 | 12252 | **12238** | 12253 |
| **Entropy** | | 0.53 | 0.57 | **0.52** | 0.58 |
| **ALMR p-value** | | <0.001 | 0.32 | **0.017** | 0.18 |
| **BLRT p-value** | | <0.001 | <0.001 | **<0.001** | <0.001 |
| **Class 1 N (%)** | 1832 (100%) | 1245 (68%) | 1200 (66%) | **563 (31%)** | 909 (50%) |
| **Class 2 N (%)** | | 587 (32%) | 274 (15%) | **321 (18%)** | 56 (3%) |
| **Class 3 N (%)** | | | 358 (20%) | **881 (48%)** | 63 (3%) |
| **Class 4 N (%)** | | | | **67 (4%)** | 547 (30%) |
| **Class 5 N (%)** | | | | | 256 (14%) |

**eTable 19** – Descriptive statistics on unimputed (available case) data.

| Diagnosis | Developmental stage | ALSPAC | | | | Pelotas | | | |
|---|---|---|---|---|---|---|---|---|---|
| | | No | Yes | Total | % Yes | No | Yes | Total | % Yes |
| Any anxiety disorder | Childhood | 7895 | 138 | 8033 | 1.7% | 3508 | 76 | 3584 | 2.1% |
| | Pre-adolescence | 7187 | 159 | 7346 | 2.2% | 3449 | 114 | 3563 | 3.2% |
| | Adolescence | 6294 | 104 | 6398 | 1.6% | 1906 | 35 | 1941 | 1.8% |
| | Young adulthood | 3726 | 419 | 4145 | 10.1 % | 2624 | 542 | 3166 | 17.1% |
| Depression | Childhood | 7928 | 52 | 7980 | 0.7% | 3568 | 16 | 3584 | 0.4% |
| | Pre-adolescence | 7479 | 74 | 7553 | 1.0% | 3549 | 14 | 3563 | 0.4% |
| | Adolescence | 6808 | 58 | 6866 | 0.8% | 1912 | 29 | 1941 | 1.5% |
| | Young adulthood | 3815 | 330 | 4145 | 8.0% | 2637 | 531 | 3168 | 16.8% |
| Specific phobia | Childhood | 8050 | 73 | 8123 | 0.9% | 3535 | 47 | 3582 | 1.3% |
| | Pre-adolescence | 7618 | 64 | 7682 | 0.8% | 3480 | 83 | 3563 | 2.3% |
| | Adolescence | 6976 | 34 | 7010 | 0.5% | 1917 | 24 | 1941 | 1.2% |
| | Young adulthood | 3995 | 150 | 4145 | 3.6% | *NA* | *NA* | *NA* | *NA* |
| Social phobia | Childhood | 8101 | 9 | 8110 | 0.1% | 3582 | <5 | 3584 | 0.1% |
| | Pre-adolescence | 7665 | 26 | 7691 | 0.3% | 3560 | <5 | 3563 | 0.1% |
| | Adolescence | 6950 | 31 | 6981 | 0.4% | 1933 | 8 | 1941 | 0.4% |
| | Young adulthood | 4056 | 89 | 4145 | 2.1% | *NA* | *NA* | *NA* | *NA* |
| GAD | Childhood | 8073 | 17 | 8090 | 0.2% | 3582 | 0 | 3582 | 0.0% |
| | Pre-adolescence | 7634 | 33 | 7667 | 0.4% | 3558 | 5 | 3563 | 0.1% |
| | Adolescence | 6933 | 31 | 6964 | 0.4% | 1937 | <5 | 1941 | 0.2% |
| | Young adulthood | 3912 | 233 | 4145 | 5.6% | *NA* | *NA* | *NA* | *NA* |
| Separation anxiety | Childhood | 8034 | 68 | 8102 | 0.8% | 3550 | 33 | 3583 | 0.9% |
| | Pre-adolescence | 7343 | 81 | 7424 | 1.1% | 3532 | 30 | 3562 | 0.8% |
| | Adolescence | 6392 | 38 | 6430 | 0.6% | 1930 | 6 | 1936 | 0.3% |
| Anxiety depression comorbidity | Childhood | 8098 | 18 | 8116 | 0.2% | 3575 | 9 | 3584 | 0.3% |
| | Pre-adolescence | 7658 | 21 | 7679 | 0.3% | 3556 | 7 | 3563 | 0.2% |
| | Adolescence | 6953 | 19 | 6972 | 0.3% | 1935 | 6 | 1941 | 0.3% |
| | Young adulthood | 3994 | 151 | 4145 | 3.6% | 3001 | 67 | 3068 | 2.2% |

#### Supplementary Figures

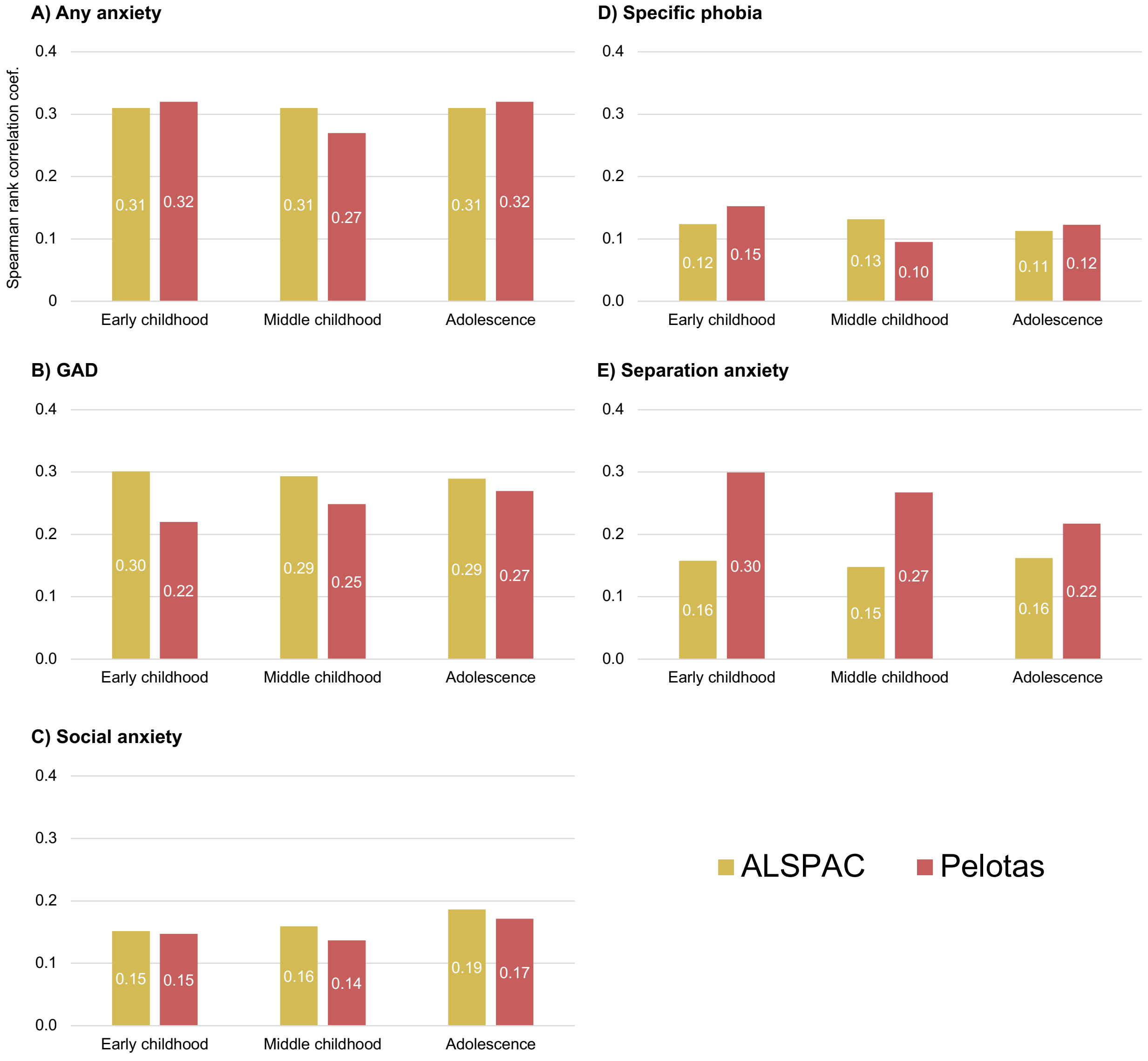


**eFigure 1 -** Correlations between any anxiety condition and depression (A), and anxiety subtypes and depression (B-E) through development in ALSPAC and Pelotas 2004 cohorts. Anxiety and depression are measured in both cohorts using the ordinal DAWBA bands.

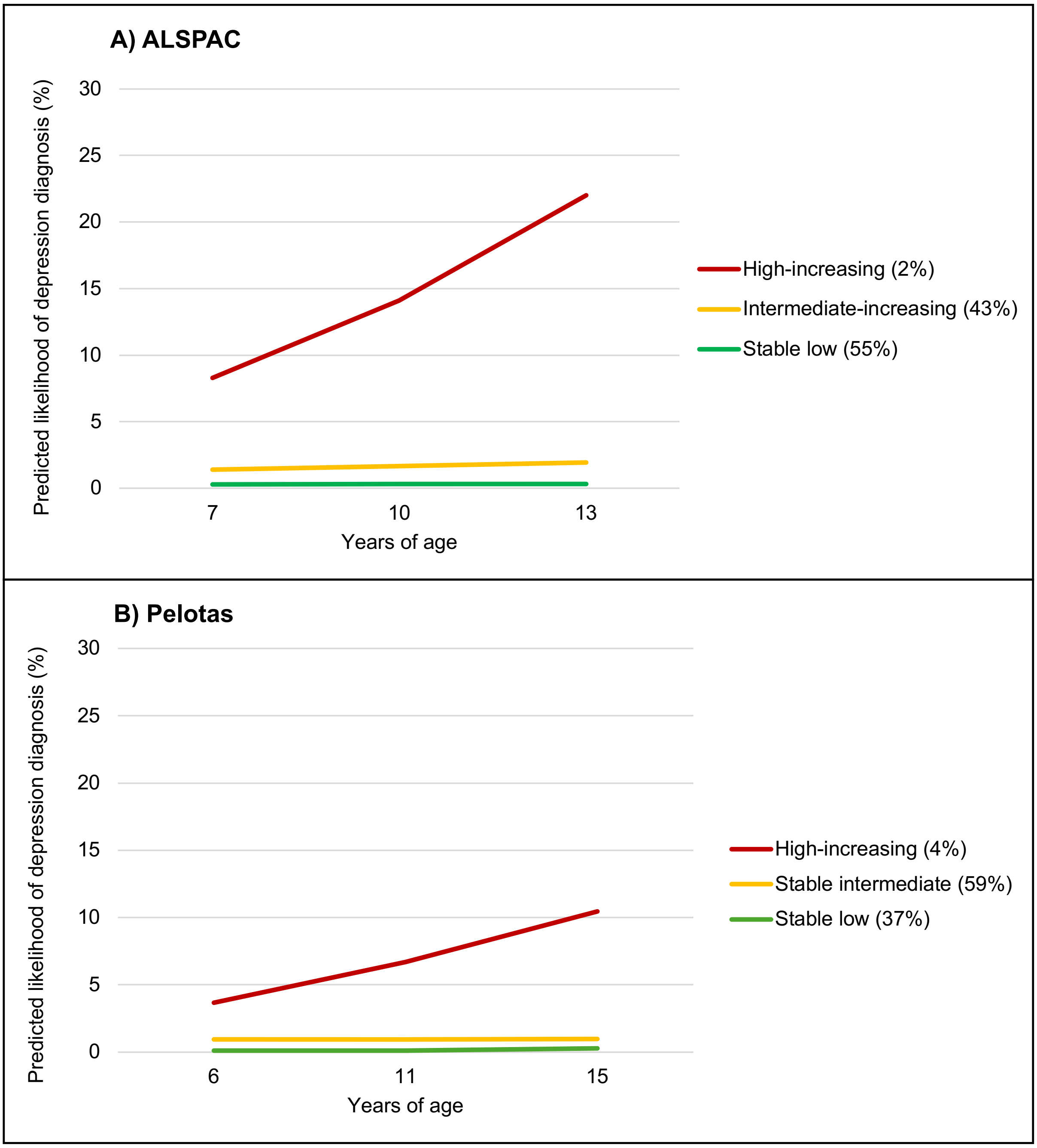


**eFigure 2** – A) Trajectories of 3-class LCGA model of depression in ALSPAC. B) Trajectories of 3-class LCGA model of depression in Pelotas 2004.

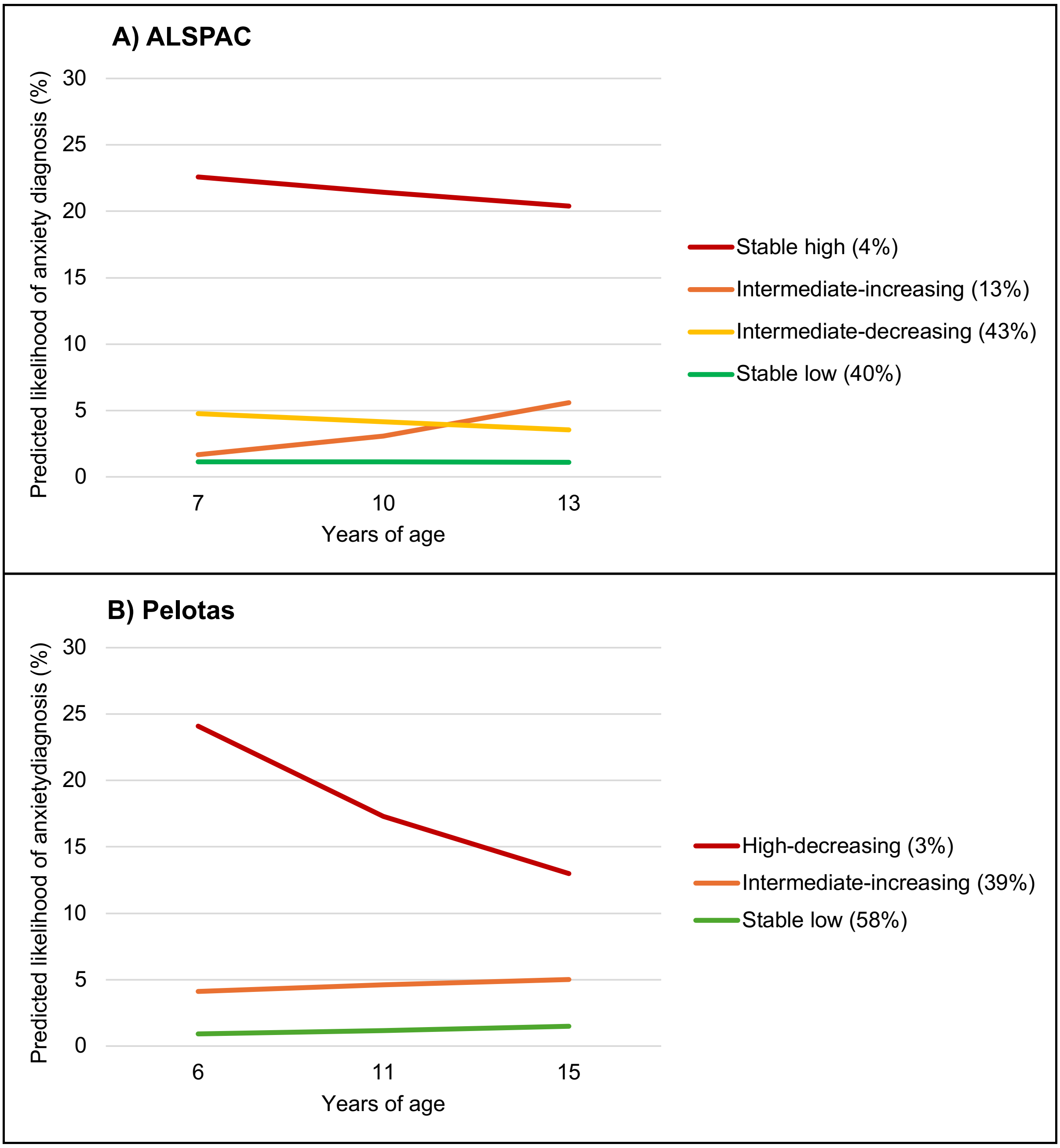


**eFigure 3** – A) Trajectories of 3-class LCGA model of anxiety in ALSPAC. B) Trajectories of 3-class LCGA model of anxiety in Pelotas 2004.

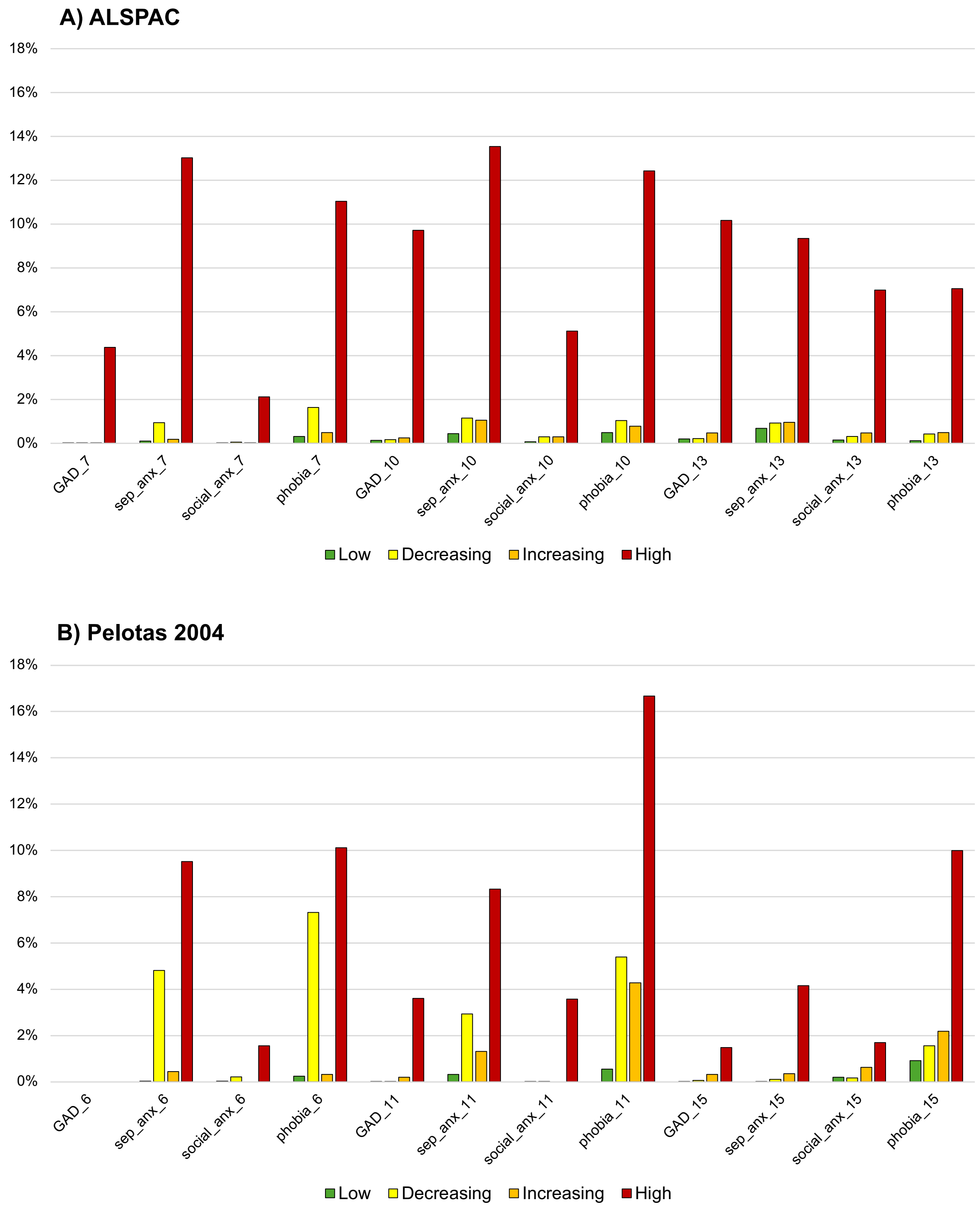


**eFigure 4 –** The prevalence of each anxiety condition in each anxiety/depression trajectory class in A) ALSPAC and B) Pelotas 2004 cohorts. For details of the trajectory groups see the main text.

#### A) ALSPAC - Males

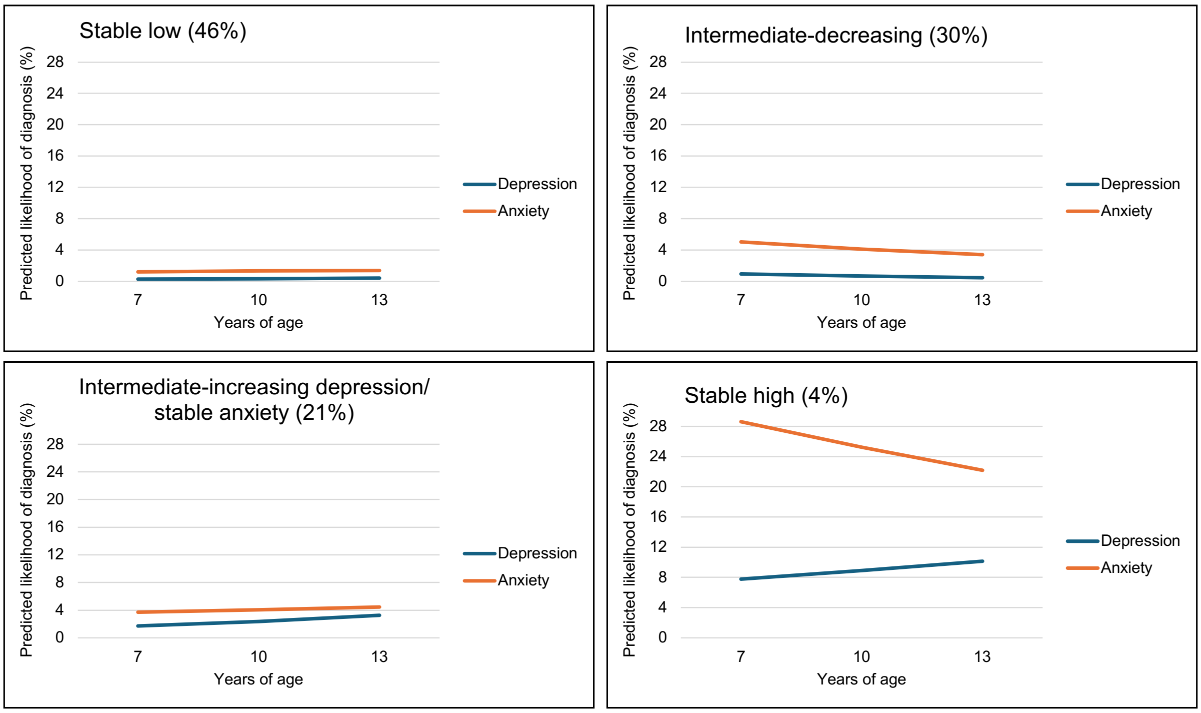


#### B) Pelotas - Males

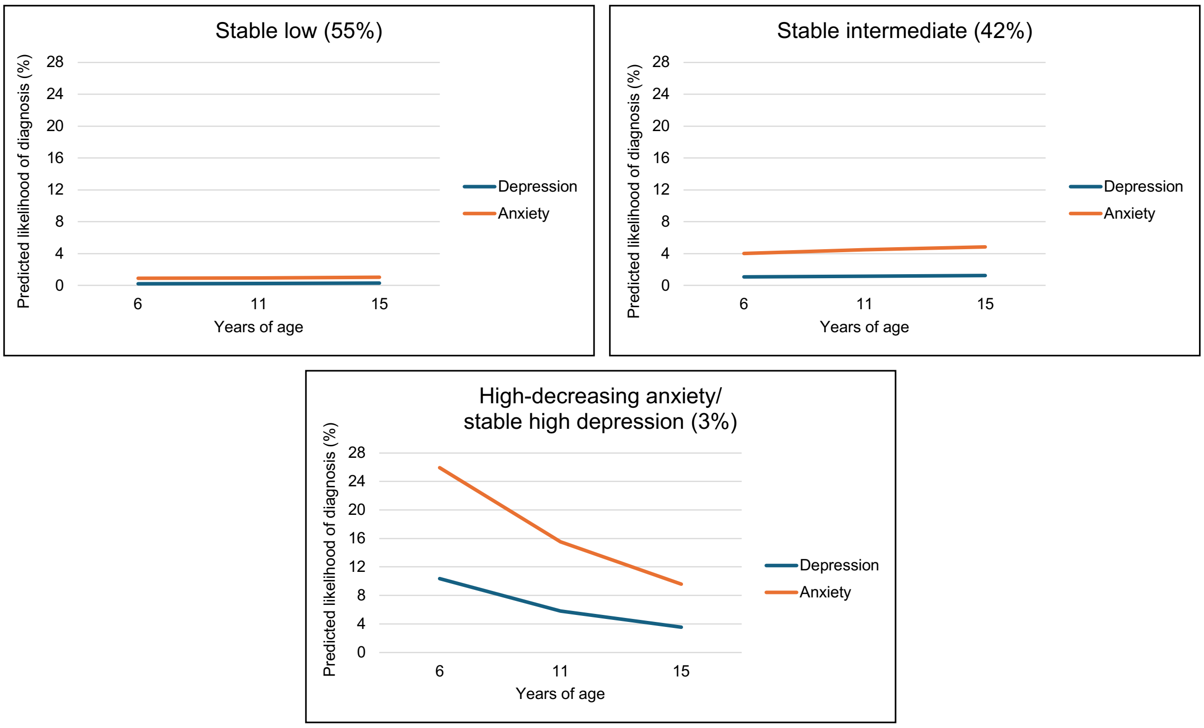


**eFigure 5** – A) Trajectories of 4-class parallel LCGA model of anxiety and depression in ALSPAC males only. B) Trajectories of 3-class parallel LCGA model of anxiety and depression in Pelotas 2004 males only.

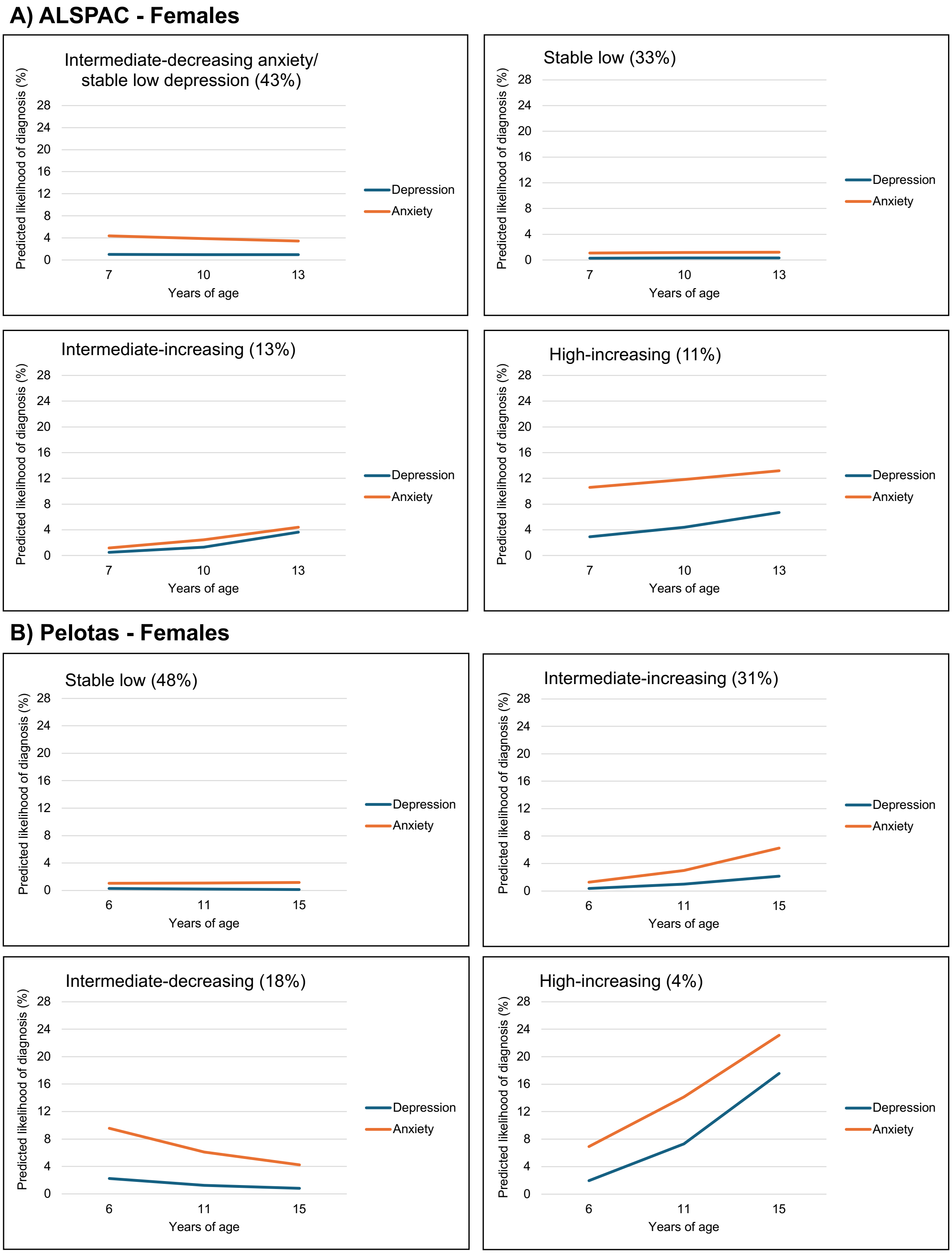


**eFigure 6** – A) Trajectories of 4-class parallel LCGA model of anxiety and depression in ALSPAC females only. B) Trajectories of 3-class parallel LCGA model of anxiety and depression in Pelotas 2004 females only.

#### Deviations from pre-registration

1. **Description of change:** DAWBA band variables (percentage likelihood of any anxiety diagnosis and depression diagnosis[1]) used for trajectories in both cohorts instead of DAWBA anxiety/depression symptom scores.
    a. **Rationale:** There were inconsistent skip rules used in ALSPAC and Pelotas 2004 cohorts, therefore symptom scores derived from individual items would not be comparable across cohorts. DAWBA band variables are comparable across cohorts.
    b. **Effect of change on study results:** The interpretation of the trajectories is slightly different, whereby rather than number of symptoms, they now reflect likelihood of diagnosis. Further, given that there is an any anxiety DAWBA band there is no need to examine the factor structure of anxiety subtype variables.

2. **Description of change:** DAWBA bands for separation anxiety, GAD, specific phobias and social anxiety were combined into an "any anxiety condition" DAWBA band, rather than performing a factor analysis across symptom scores for anxiety conditions.
    a. **Rationale:** DAWBA anxiety/depression symptom scores were not used (see above).
    b. **Effect of change on study results:** A factor analysis would have allowed assessment of whether individual anxiety conditions load onto a common latent anxiety construct. By using the composite "any anxiety condition" DAWBA band, we were unable to examine the underlying factor structure.

3. **Description of change:** Growth curve modelling used to examine the relationship between anxiety and depression across development, as well as cross-sectional correlations across time.
    a. **Rationale:** Using growth curve modelling allows interpretation of relationship between intercept (baseline) and slope (rate of change) of anxiety and depression, while accounting for time-specific residual correlations between anxiety and depression.
    b. **Effect of change on study results:** N/A.

**Analysis scripts**

1. Parallel-process growth curve modelling of anxiety and depression

```
DATA:
  FILE = [];
VARIABLE:
  NAMES = dep7 anx7 dep10 anx10 dep13 anx13;
  USEVAR = dep7 anx7 dep10 anx10 dep13 anx13;
  CATEGORICAL = dep7 anx7 dep10 anx10 dep13 anx13;
  MISSING=.;
 ANALYSIS:
   MODEL = NOCOV;
 MODEL:
   I_D S_D | dep7@0 dep10@3 dep13@6;
   I_A S_A | anx7@0 anx10@3 anx13@6;

   dep7 WITH anx7;
   dep10 WITH anx10;
   dep13 WITH anx13;

! These are included/removed as necessary to improve model fit
   I_A WITH I_D;
   S_A WITH S_D;
   I_A WITH S_A;
   I_D WITH S_D;
   I_D WITH S_A;
   I_A WITH S_D;

  OUTPUT: SAMPSTAT STANDARDIZED MOD (5) res;
```

2. Parallel-process 2-class LCGA of anxiety and depression

```
DATA:
  FILE = [];
VARIABLE:
  NAMES = dep7 anx7 dep10 anx10 dep13 anx13;
  USEVAR = dep7 anx7 dep10 anx10 dep13 anx13;
  CATEGORICAL = dep7 anx7 dep10 anx10 dep13 anx13;
  MISSING=.;
  CLASS = c (2); !change based on class N
ANALYSIS:
  TYPE = MIXTURE;
  STARTS = 500 10;
  LINK = LOGIT;
MODEL:
 %OVERALL%
   I_D S_D | dep7@0 dep10@3 dep13@6;
   I_A S_A | anx7@0 anx10@3 anx13@6;
OUTPUT: res TECH7 TECH11 TECH14;
```

3. Bias-adjusted 3-step association of trajectory classes with outcomes

```
DATA:
      FILE = [];
      TYPE = IMPUTATION;
VARIABLE:
      NAMES = class MDD_18_CISR Anx_18_CISR;
      USEVAR = class MDD_18_CISR;
      NOMINAL = class;
      MISSING=.;
      CLASS = c(4);
ANALYSIS:
      TYPE = MIXTURE;
MODEL:
  %OVERALL%
  c ON MDD_18_CISR;
   %c#1%
   [class#1@8.190 class#2@5.387 class#3@5.966];
  %c#2%
   [class#1@1.533 class#2@3.325 class#3@1.873];
  %c#3%
   [class#1@2.312 class#2@2.722 class#3@3.595];
   %c#4%
   [class#1@-6.062 class#2@-0.846 class#3@-2.808];
```
